# Association between time spent in the emergency department and 30-day mortality: a population-level observational study in England

**DOI:** 10.1101/2025.06.09.25329259

**Authors:** Hannah Aston, Perrine Machuel, Nina Mill, Owen Gethings, Benjamin Bloom, Adrian Boyle, Ian Higginson, Chris Moulton, Janos Suto, Vahé Nafilyan, Daniel Ayoubkhani

## Abstract

**Objective:** To investigate the association between patient time spent in Type 1 emergency departments (EDs) and all-cause mortality 30 days after leaving the department alive.

**Design:** Cross-sectional, retrospective, observational study using national linked data.

**Setting:** All NHS Type 1 emergency departments in England.

**Participants:** 6,721,179 individuals (mean age 41.3 years, 52.6% female, 81.4% White ethnicity) attended an ED at least once between 21st March 2021 and 31st March 2022; had a record populated with a “non-immediate” acuity level and a chief complaint at arrival; and survived to either discharge from the ED or admission to hospital for inpatient care.

**Main outcome measures:** All-cause mortality within 30 days of the ED attendance.

**Results:** Within the study population, 88,657 patients (1.3%) died within 30 days of ED attendance. A positive non-linear relationship was observed between time spent in the ED and post-discharge mortality, with the probability of death increasing after two hours. The marginal probabilities of death (controlling for socio-economic characteristics, clinical factors such as chief complaint, and comorbidities) at two hours in the ED were 0.02% for patients aged 20 years, rising to 0.1% at 40 years, 0.3% at 60 years, and 0.8% at 80 years. Compared with patients who spent two hours in the ED, the adjusted odds of post-discharge death were: 1.1 times higher (1.07 to 1.14) for three hours; 1.6 times (1.48 to 1.68) for six hours; 1.9 times (1.80 to 2.03) for nine hours; and 2.1 times (2.02 to 2.28) for 12 hours.

**Conclusions:** Longer time spent in the ED for non-immediate care is associated with increased risk of all-cause mortality within 30 days of discharge or admission, in a non-linear manner. Our findings suggest that time in the ED may be a risk factor for death after discharge, not just during the visit. These findings could inform policy makers and health professionals when setting ED time targets. Further research is needed to understand causal drivers of post-discharge mortality and confirm whether our findings generalise to more recent periods.

**Summary box:** *Section 1: What is already known on this topic:*

- Small, single-centre studies have suggested that there is an increased mortality rate among patients who experience delays between arrival at the emergency department and admission to an inpatient bed.
- A larger study of over five million individual admitted patients in England, using data from between 2016 and 2018, found an increase in all-cause 30-day mortality that was associated with delays to hospital admission. The quantifiable increase in mortality started as early as five hours after arrival at the emergency department (ED) and increased in a linear “dose-related” fashion.

*Section 2: What this study adds:*

- Our study suggests that longer ED stays for non-immediate care are associated with an increased risk of all-cause mortality within 30 days of both discharge or admission.
- Our study builds on previous research by incorporating more recent data, treating time spent in the ED as a continuous rather than a discrete variable; controlling for a broad range of socio-economic variables recorded in England and Wales National Census; and including all patients who attended emergency departments, for both those that were admitted and those that were discharged (rather than just those who were subsequently admitted to hospital for inpatient care).

## Introduction

Over the past decade, the waiting time for emergency department (ED) patients in England has increased substantially [1]. Changes in technology and clinical practice, along with increases in population size, changes in demographic composition and a reduction in acute hospital bed capacity have contributed to crowding in EDs and delays for patient care throughout the UK [2, 3]. Time spent waiting for emergency admission to inpatient care has been impacted by high bed occupancy rates [4, 5], delays in transferring individuals out of hospital due to pressures on other clinical and social services [6], and staff shortages [7].

In 2004, the National Health Service (NHS) introduced a four-hour operational standard, by which 95% of patients should be discharged, admitted or transferred within four hours of arrival at an ED. Additional pressures in recent years, such as the Coronavirus Disease 2019 (COVID-19) pandemic, have led to further increases in time spent in emergency care, with the majority of EDs consistently missing the four-hour target [8].

Previous studies have demonstrated that increased waiting times for emergency and critical care are associated with higher mortality rates after leaving ED care [9–12]. In the largest study to date, delays to inpatient admission in excess of five hours were found to be associated with an increase in all-cause mortality within 30 days of leaving the ED, and a 10% increase in post-discharge mortality for patients remaining in the ED for 12 hours when compared with those who leave after six hours [9]. Similarly, both time to be seen by a clinician and time from being seen to inpatient admission have been found to be strong predictors of death within 30 days of departure [10]. Smaller scale, single-centre studies in Canberra, Australia [11] and Washington, USA [12] have also found associations between waiting time and mortality. Though insightful, these studies were limited by small sample sizes, the use of binary or coarse time intervals for the waiting time exposure, and the fact that most are now over 10 years old.

Notably, time spent in EDs has increased substantially in the UK over this period, and the COVID-19 pandemic led to changes to operational procedures in EDs and contributed to overall population mortality rates. There may have also been changes to the case mix and underlying acuity level of people attending EDs. It is therefore unclear if these studies are generalisable to more recent periods.

In this study, we investigated the association between time spent in Type 1 EDs (consultant led 24-hour service with full resuscitation facilities and designated accommodation for the reception of accident and emergency patients) and all-cause mortality 30 days after leaving the department alive, using newly linked 2021 Census data and electronic health records covering most of the population of England. Unlike previous studies, we were able to account for a wide range of socio-economic factors, comorbidities and indicators of illness severity at arrival, and model time spent in the ED in a continuous manner, using a contemporary dataset with a large, near-complete study population.

## Methods

### Study design and data sources

We conducted a cross-sectional, retrospective, observational study of all individuals who attended a Type 1 ED in England between 21st March 2021 (Census day) and 31^st^ March 2022 (the last ED records available to us at the time of analysis).

We created a person-level dataset combining NHS England’s Emergency Care Dataset (ECDS) on ED attendances with: the 2021 Census; the Hospital Episode Statistics (HES) Admitted Patient Care (APC) and Outpatient (OP) datasets; and Office for National Statistics (ONS) death registration data. The 2021 Census was linked to the Personal Demographics Service (PDS) to retrieve NHS numbers of Census respondents, with a linkage rate of 95.8% [13]. All other datasets were subsequently linked to the Census using NHS numbers.

ECDS [14] is the national dataset for urgent and emergency care and replaced the Accident and Emergency Commissioning Data Set in 2019. It provides information to support the care provided in EDs and includes data items needed to understand a patient’s journeys in the ED. Based on the approach of Jones et al. (2022) [9], we only included the individuals’ first ED attendance during the study period in order to maintain the independence of observations in the study sample.

HES [15] contains records of all individuals treated in NHS hospitals in England, covering details of inpatient admissions and outpatient appointments.

Census 2021 [16] contains records of all people and households in England and Wales and information about that population such as socio-demographic and socio-economic characteristics, collected via a questionnaire (completed online for most people). The estimated response rate was 97% of the usual resident population of England and Wales, and more than 88% in all local authorities.

### Study population

Our study population included individuals who were enumerated at the 2021 Census as usual residents of England who: could be linked to the 2019 NHS PDS; visited a Type 1 ED at least once between 21^st^ March 2021 and 31^st^ March 2022; had an ECDS record that was populated with a “non-immediate” acuity level (“low”, “standard”, “urgent”, or “very urgent”) and a chief complaint (reason for attending the ED) at arrival; and who survived to discharge from the ED or admission to hospital for inpatient care. The analysis only covered patients with non-immediate acuity levels (all acuity levels other than “immediate”), who represent 98.5% of the total number of ED patients. For our main analysis, we aggregated the non-immediate acuity groups in this way to increase statistical power. In addition, there is likely to be a degree of inconsistency in how acuity levels are coded in different EDs (i.e. measurement error). The 1.5% of patients in the “immediate” acuity group (those requiring resuscitation on arrival) were excluded from the analysis because of the high mortality rate within a short time after arriving at the ED in this group, a confounding factor that we could not account for in our analysis and which would therefore introduce bias if we retained this group in the study population (See Supplementary Figure 1).

We excluded individuals recorded as deceased on arrival and removed records that we deemed unreliable: departure date occurring before arrival date; missing information on arrival or departure date or location; missing information on method of arrival and admittance; or recorded time in the ED exceeding 48 hours.

### Exposure

We measured time spent in the ED in terms of patients’ end-to-end length of stay (the difference in hours and minutes between the arrival and departure times recorded in ECDS).

### Outcome

The primary outcome was all-cause mortality within 30 days of an ED attendance, after leaving the department alive, for deaths occurring between 21^st^ March 2021 (start of study period) and 30^th^ of April 2022 (30 days after the last ED record available to us).

### Covariates

We adjusted for a range of covariates recorded on the ECDS dataset hypothesised to be related to both time spent in the ED and all-cause mortality: age at arrival to the ED, arrival via emergency ambulance, month of arrival, time of day of arrival and reason for attendance (recorded as chief complaint).

Covariates recorded in Census 2021 were: sex; ethnicity; region of residence; local area deprivation decile group (Index of Multiple Deprivation [17]); highest qualification; eight categories of the National Statistics Socio-Economic Status [18] (NS-SEC); self-reported long-term health condition or disability status, and self-reported general health.

Comorbidities were derived from HES APC and OP data over the five years prior to 21 March 2021 as binary flags indicating whether the individual had received hospital treatment for each condition. The comorbidities included were: COVID-19, cancer, diabetes, dementia, serious mental illness, autism, neurological motor neuron disease and Parkinson’s Disease and Multiple Sclerosis (MS), Alzheimer’s disease, epilepsy, hypertension, angina, myocardial infarction, ischaemic heart disease, atrial fibrillation, heart failure, stroke, influenza or pneumonia, other respiratory infections, chronic obstructive pulmonary disease (COPD) or respiratory failure, asthma, inflammatory bowel disease, liver disease, rheumatoid arthritis, osteoarthritis, and kidney disease.

A full description of the variables included in analysis can be found in Supplementary Table 1.

### Statistical analysis

We described the characteristics of the study population using means and standard deviations for age and counts and proportions for all discrete covariates. We compared the covariate distributions according to whether individuals spent less than four hours versus at least four hours in ED using standardised differences, with absolute values greater than 10% indicating large imbalance between covariate distributions [19].

We fitted a logistic regression model to assess the association between time spent in the ED and the risk of all-cause, 30-day death following departure. We modelled age at ED arrival and hours spent in the ED using restricted cubic splines [20], also known as “natural splines”, by fitting a set of unique curves (piecewise polynomials) between pre-defined percentiles (“knots”) of the distribution of each variable. The restricted cubic spline enforces the relationship between each variable and mortality to be linear beyond the outermost knots to avoid the distorting effect of extreme values. We fitted a series of models with bespoke knot combinations for age and time spent in the ED. Our final model uses the knot combination that minimised the value of the Bayesian Information Criterion (BIC) (see Supplementary Table 2). All variables other than age at arrival and time spent in the ED were modelled discretely using reference coding.

The model was used to estimate marginal probabilities of 30-day all-cause mortality as a function of time spent in the ED using R’s emmeans package [21]. These estimates represent the probability of death at each unit of time spent in ED, averaged over each combination of covariates included in the model (a weighted average, as per the observed frequencies in the study population). The odds ratios for mortality risk at different times spent in the ED were referenced to the risk at two hours (the nadir of the fitted curve). All point estimates were accompanied by 95% confidence intervals (CIs).

We tested for interactions between time spent in the ED and several potential effect-modifiers: age, sex, region, whether the patient was admitted to hospital or discharged to home (to build on the results of Jones et al (2022) [9]), relative area deprivation decile group, and chief complaint at arrival. Interactions with p<0.05 (based on a likelihood ratio test compared with the model without the interaction term) were deemed to be statistically significant.

### Sensitivity analysis

We conducted sensitivity analysis to evaluate the robustness of our results to the inclusion of additional covariates. We adjusted for admission status (admitted or discharged) as sensitivity analysis and not in the main model as admission status is assigned at the end of the ED visit rather than at the start, and therefore it may be viewed as a post-treatment variable and not appropriate to include in the main analysis. In our main analysis, we aggregated the non-immediate acuity groups together. As sensitivity analysis, we produced results for each individual group (see Supplementary Figure 2).

All data preparation was conducted using Sparklyr version 1.8.6 and PySpark version 3.8. All statistical analyses were performed using R version 3.5.1.

## Patient and public involvement

Due to the general nature of our study population (all ED attendances for any reason, rather than with specific health conditions) and outcome definition (mortality), we did not directly involve patients and the public in the study.

## Ethical approval

This study was ethically self-assessed against the ethical principles of the National Statistician’s Data Ethics Advisory Committee (NSDEC) using NSDEC’s ethics self-assessment tool. We subsequently engaged with the UK Statistics Authority Data Ethics team, who were satisfied that no further ethical approval was required.

## Results

### Characteristics of the study population

Between 21^st^ March 2021 and 31^st^ March 2022, 6,721,179 individuals attended an ED and met our study inclusion criteria. See Supplementary Figure 3 for the sample flow diagram. The mean age of the study population was 41.3 years, 52.6% of individuals were female and 81.4% were White.

Compared with individuals who spent <4 hours in the ED, Table 1 shows that patients who spent ≥4 hours in the ED were more likely to: have arrived between 6am to 12pm or 12pm to 6pm (67.3% vs. 54.0%); live in the North East, West Midlands, Yorkshire and the Humber, or London (44.1% vs. 40.0%); live in the 30% most deprived areas (35.4% vs. 23.7%); have a self-reported health status of “very good” or “good” (77.4% vs. 57.5%); have no long-term health conditions/disabilities (71.0% vs. 53.9%); be attending the ED for one of the following reasons: “eye”, “head and neck”, “obstetrics and gynaecology”, “skin” and “trauma / musculoskeletal” (53.5% vs. 28.7%); not arrive at the ED via emergency ambulance (83.8% vs. 58.7%); and not be admitted to hospital at the end of the ED visit (83.6% vs. 58.8%).

**Table 1.**
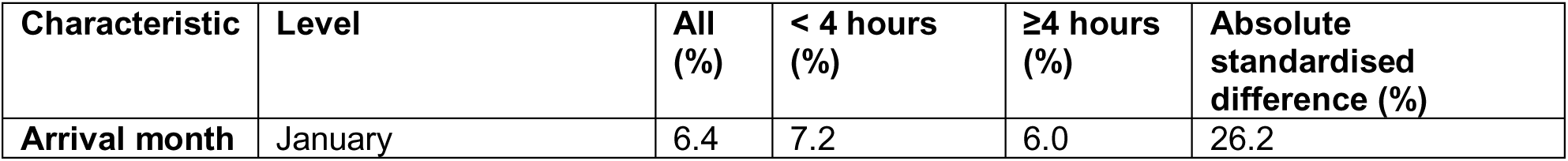
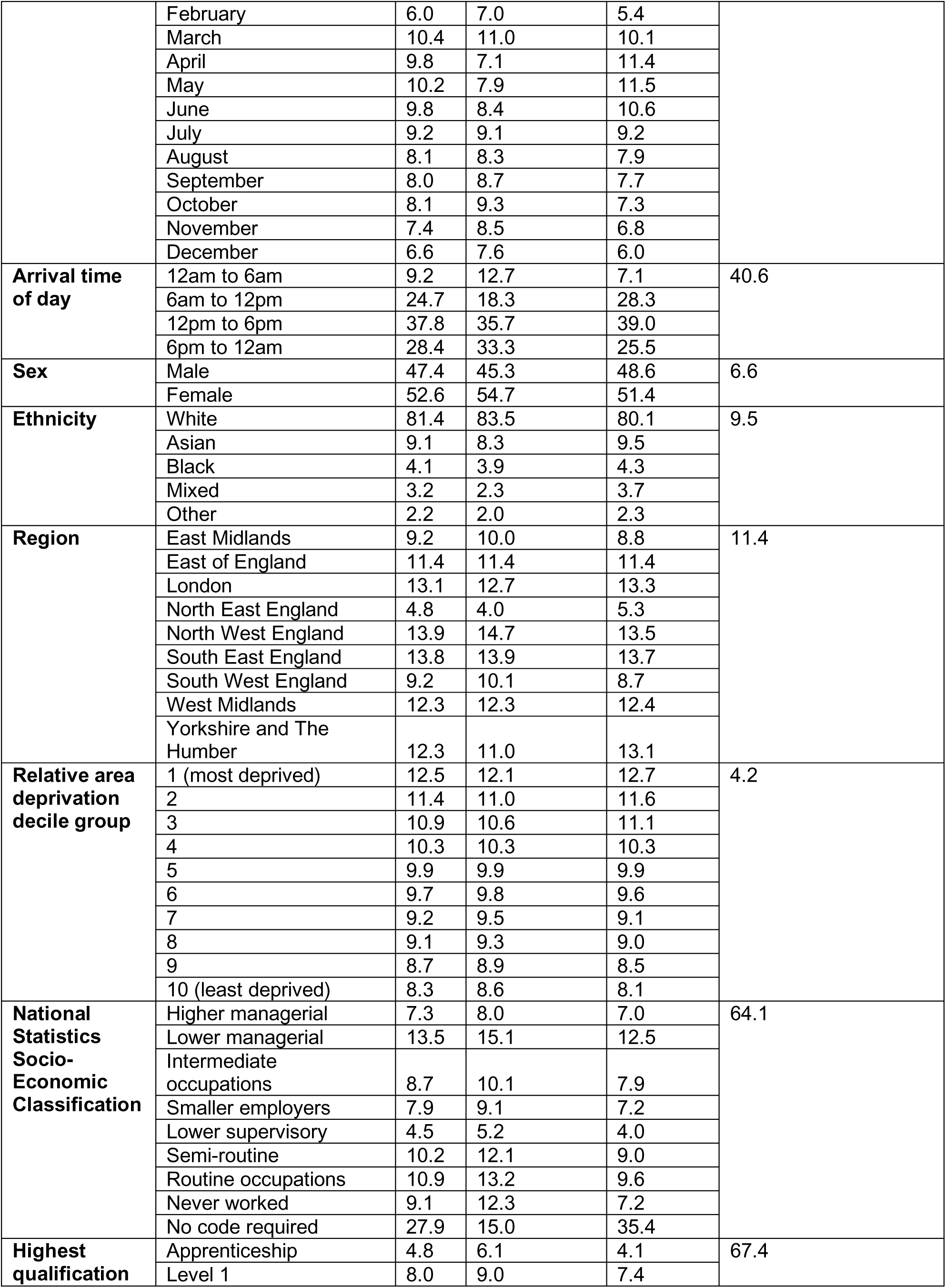
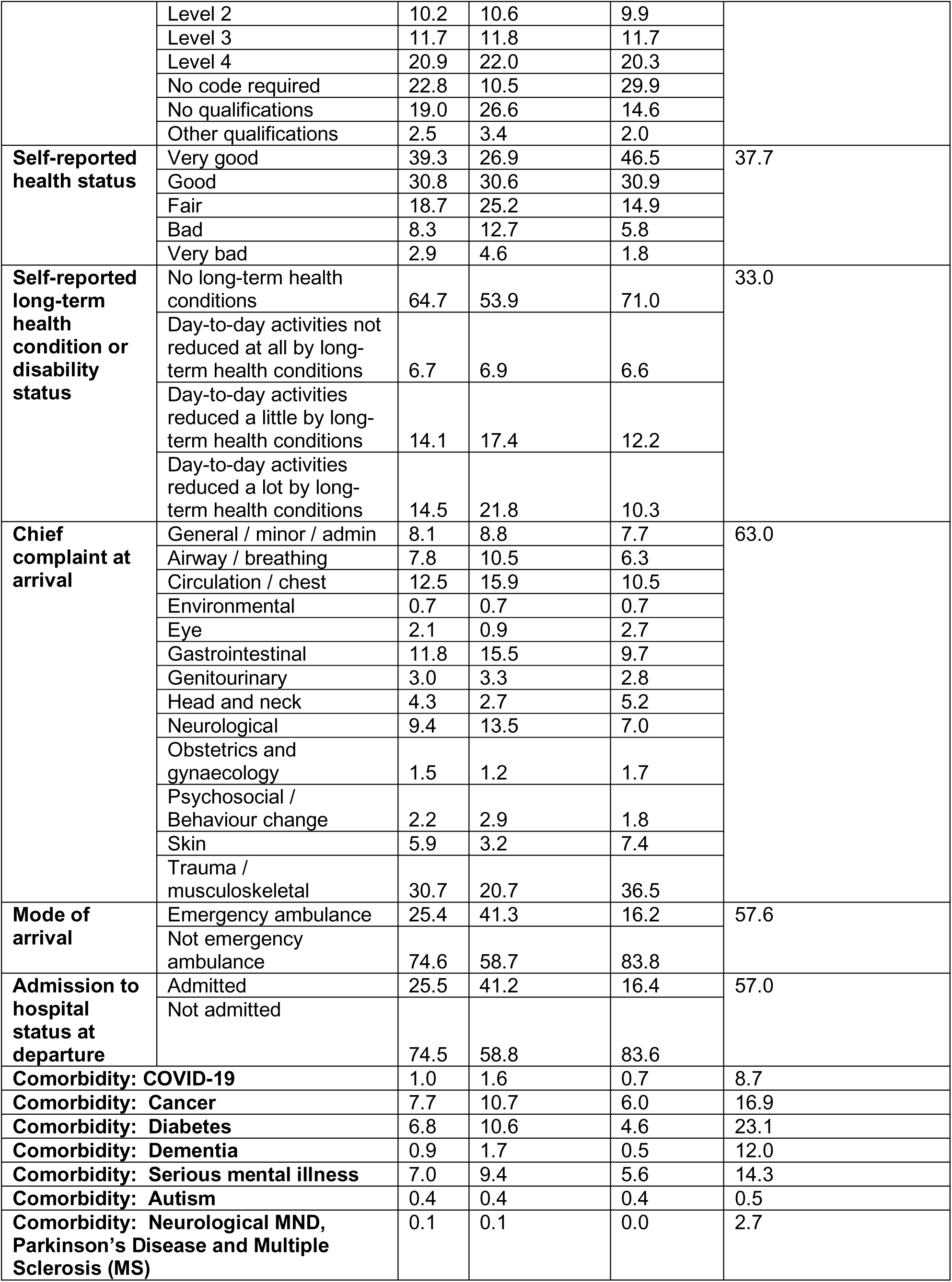
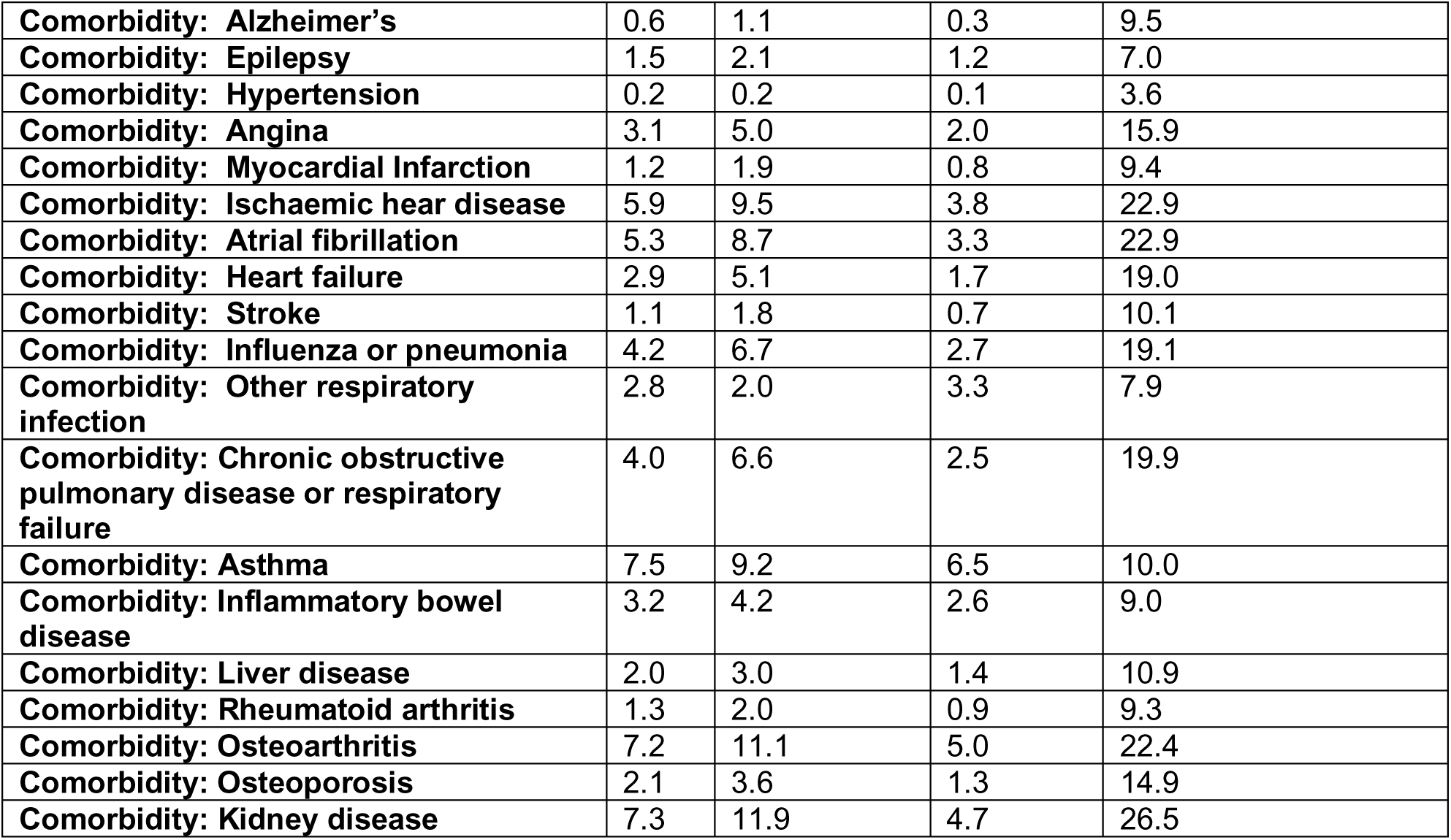
Distributions of covariates by time spent in the ED (<4 hours versus ≥4 hours)

Median time spent in the ED was 3.6 hours, interquartile range 2.3 to 5.5 hours. 70.8% of patients were in the ED for more than two hours, 32.8% for more than four hours, and 4.2% for more than 12 hours. 88,657 individuals (1.3%) died within 30 days of their ED attendance. Time spent in the ED is not normally distributed, with a sharp discontinuity in the distribution at four hours (Figure 1).

**Figure 1.**
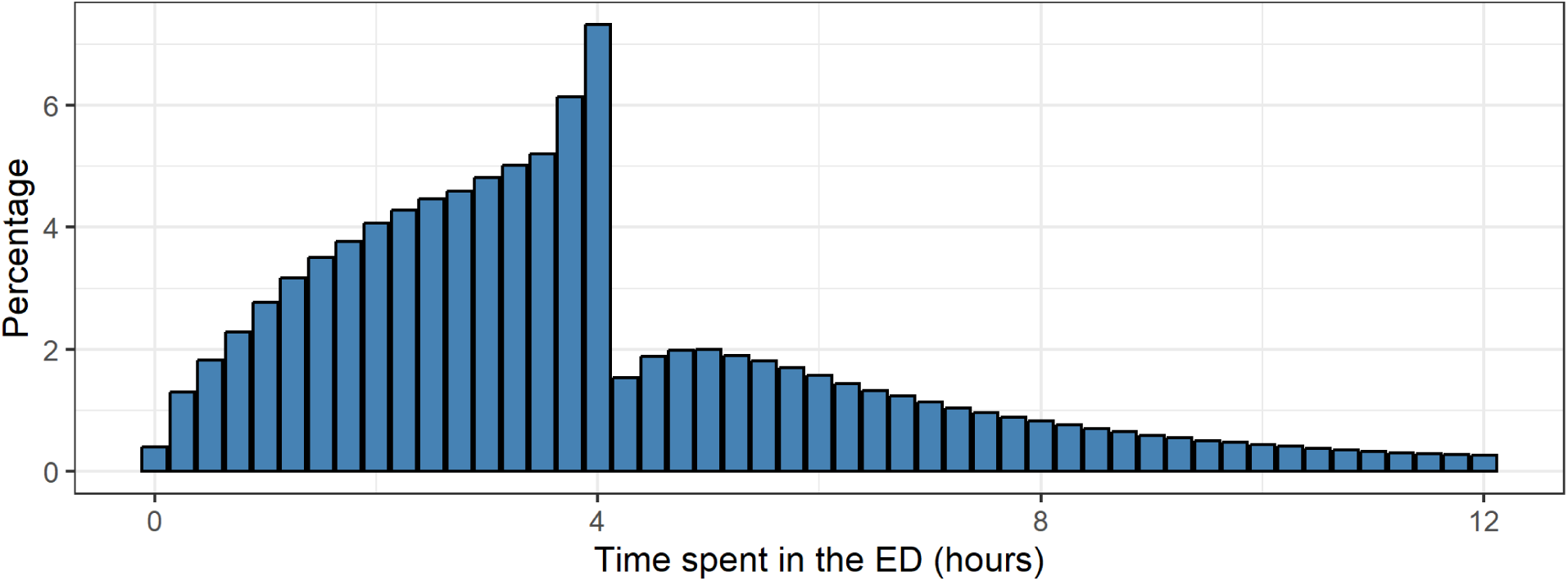
Distribution of total time spent in ED Notes: 1. The chart has been truncated at 12 hours. Counts up to 48 hours can be found in Supplementary Figure 4. 2. Time spent in the ED has been rounded to the nearest hour.

### Adjusted absolute risk of 30-day post-discharge mortality

The estimated marginal probability of dying within 30 days of discharge from the ED increased with age in a non-linear manner (Figure 2). At two hours of total time spent in the ED (from arrival to discharge), 0.02% of patients aged 20 years died post-discharge, increasing to 0.1% for patients aged 40 years, 0.3% for patients aged 60 years, and 0.8% for patients aged 80 years.

**Figure 2.**
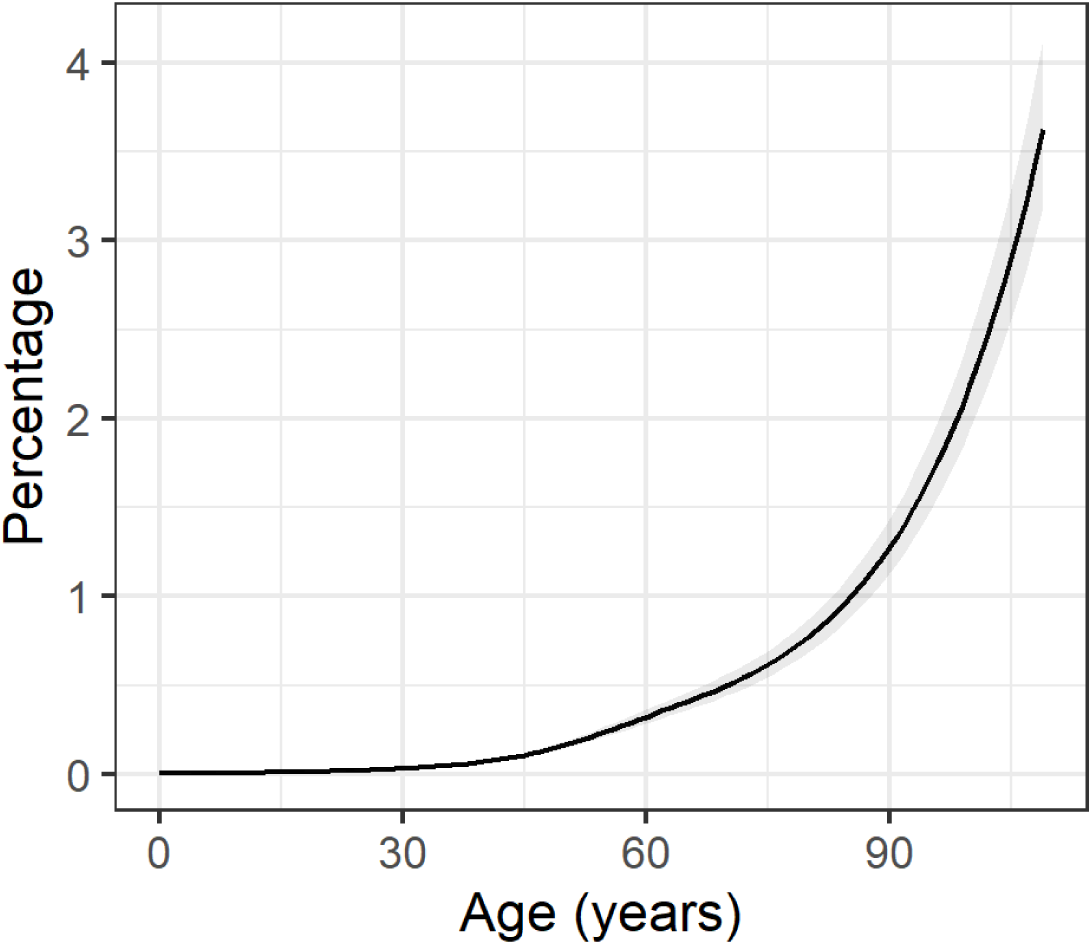
Estimated marginal percentage of ED patients who died within 30 days of discharge by age, at two hours spent in the department Notes: 1. These are modelled estimates, marginalised over the covariates outlined in the Methods section. The estimates are standardised to the covariate distributions observed in the study population. 2. The shaded area represents the 95% CI around the point estimates.

For counts of how many patients died by time spent in the ED and age, sex, region of residence, admission status, relative area deprivation decile group, chief complaint and acuity level, see Supplementary Tables 3 to 10.

### Association between time spent in ED and 30-day post-discharge mortality

After adjusting for the confounding factors outlined in the Methods section, we found that the risk of post-discharge death increased with total time spent in the ED after approximately two hours (Figure 3). Compared with patients who spent two hours in the ED, the odds of post-discharge death were: 1.1 times higher (95% CI: 1.07 to 1.14) for those who spent three hours in the ED; 1.6 times higher (1.48 to 1.68) for those who spent six hours in the ED; 1.9 times higher (1.80 to 2.03) for those who spent nine hours in the ED; and 2.1 times higher (2.02 to 2.28) for those who spent 12 hours in the ED. For patients spending longer than 12 hours in the ED, when compared to patients who spent two hours, the odds of post-discharge death were: 2.4 (95% CI: 2.2 to 2.6) times higher for those who spent 24 hours in the ED; and 2.6 (2.2 to 3.2) times higher for those who spent 48 hours in the ED. Estimates up to 48 hours can be found in Supplementary Figure 5.

**Figure 3.**
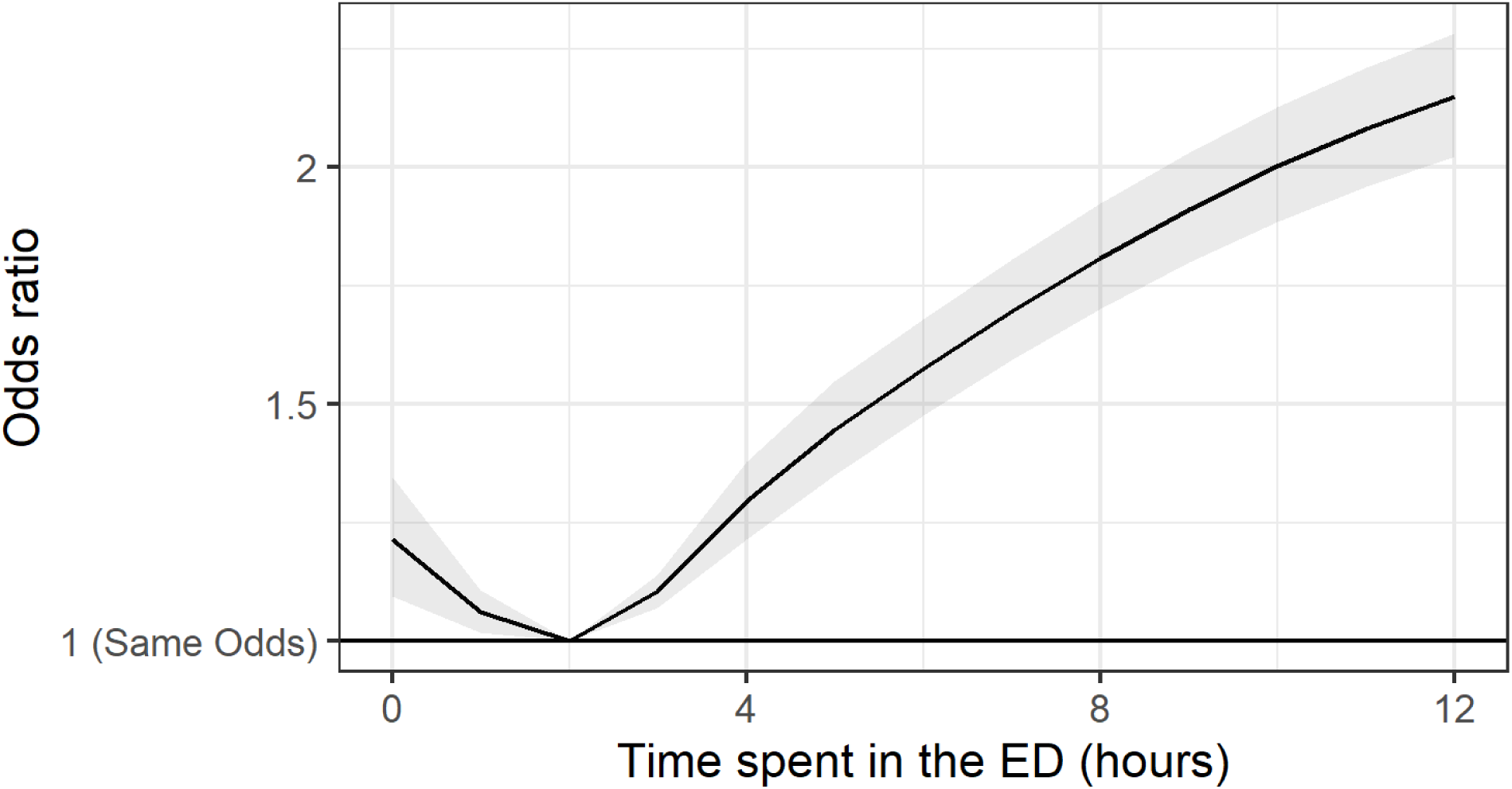
Adjusted odds ratios for all-cause, 30-day mortality as a function of time spent in the ED, compared to two hours Notes: 1. The estimates are adjusted for the covariates outlined in the Methods section. 2. The chart has been truncated at 12 hours. Estimates up to 48 hours can be found in Supplementary Figure 5. 3. The shaded area represents the 95% CI around the point estimates.

### Effect-modifiers

Age (p<0.001), region of residence (p<0.001), admission status at departure (p<0.001), relative area deprivation decile group (p<0.001), chief complaint at arrival (p<0.001) and acuity level (p<0.001) were all found be significant modifiers of the relationship between time spent in the ED and post-discharge mortality risk. The interaction between time spent in the ED and sex was not significant at the 5% level (p=0.37).

The odds ratio for post-discharge mortality after 12 hours in the ED compared with two hours was greatest for younger patients. For example, the odds at 12 hours were 4.6 (95% CI: 2.01 to 10.37) times higher than at two hours for patients aged 20 years old. See Supplementary Figure 6.

Individuals living in London had the highest regional odds ratio for post-discharge mortality after 12 hours spent in the ED compared with two hours, with odds 4.6 (95% CI: 2.07 to 3.43) times higher. Individuals living in the South West of England had the smallest odds ratio for mortality after 12 hours compared to two hours, with odds 1.8 times (95% CI: 1.49 to 2.10) higher. See Supplementary Figure 7.

Individuals who were not admitted to hospital at the end of their ED visit had a higher odds ratio than admitted patients for post-discharge mortality after 12 hours spent in the ED compared with two hours (Figure 4), with odds 2.8 (95% CI: 2.49 to 3.19) times higher. Conversely, the odds for patients who were admitted to hospital and spent 12 hours in the ED were 1.4 (95% CI: 1.31 to 1.52) times higher than admitted patients who spent two hours in the ED.

**Figure 4.**
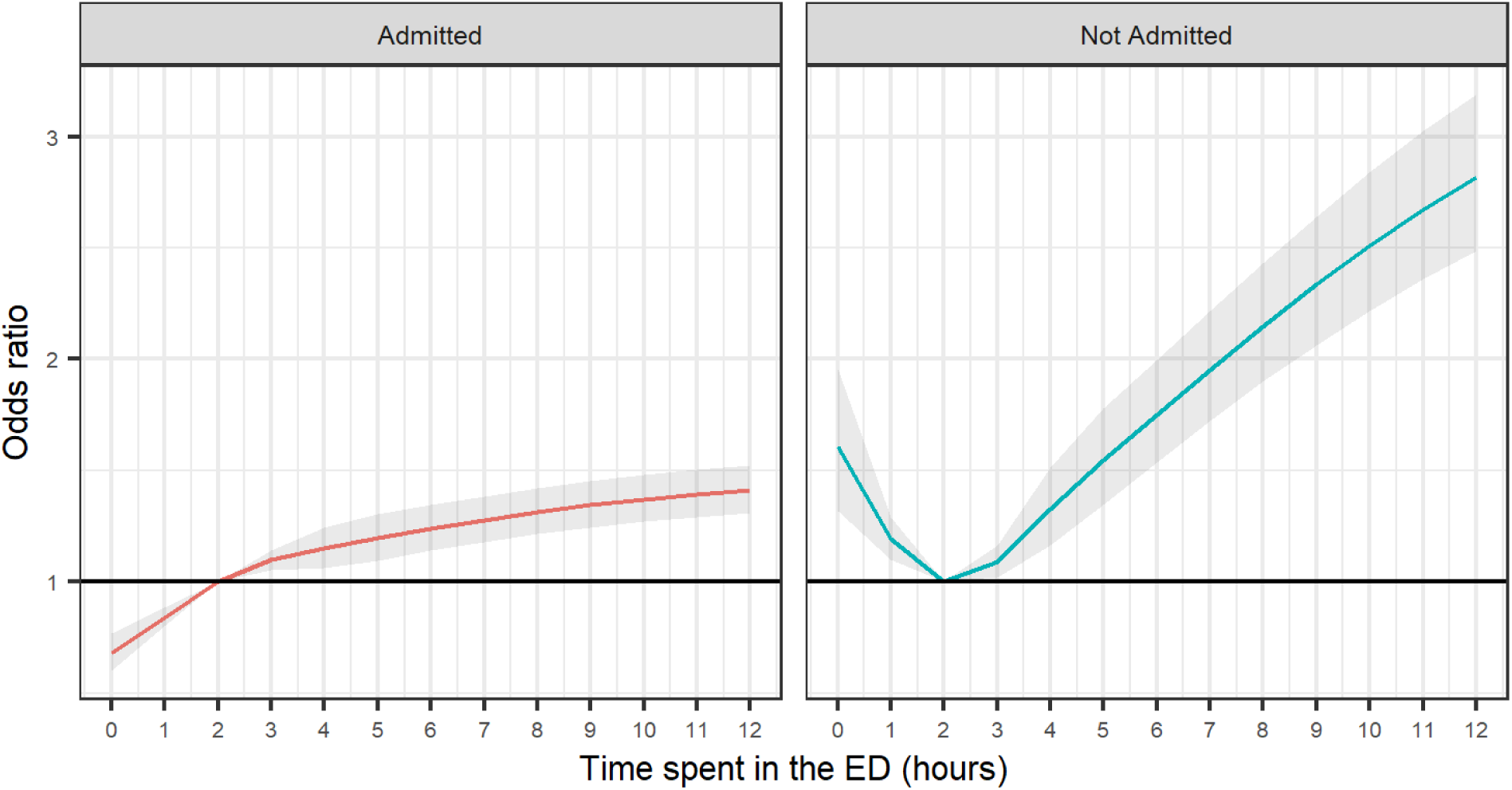
Odds ratios for all-cause, 30-day mortality as a function of time spent in the ED, by hospital admission status, compared with two hours Notes: 1. The estimates are adjusted for the covariates outlined in the Methods section. 2. The shaded area represents the 95% CI around the point estimates.

The odds ratio for post-discharge 30-day mortality after 12 hours in the ED compared with two hours was highest for individuals living in area deprivation decile group 3 (i.e. people living in the third 10% most deprived areas of England), with odds 2.5 (95% CI: 2.04 to 2.99) times higher. Individuals living in area deprivation decile group 8 had the lowest odds ratio for mortality at 12 hours spent in the ED when compared to two hours, with odds 2.0 (95% CI:1.64 to 2.34) times higher. See Supplementary Figure 8.

Individuals attending an ED who were recorded as having problems with their eyes had the highest odds ratio for mortality after 12 hours spent in the ED when compared to two hours (Figure 5), with odds 7.9 (95% CI: 2.28 to 27.69) times higher. Conversely, patients attending an ED for neurological reasons had the lowest odds ratio for mortality after 12 hours spent in the ED when compared to two hours, with odds 1.4 (95% CI: 1.27 to 1.62) times higher. However, these results should be interpreted cautiously due to the relatively wide CIs around the point estimates for some chief complaints, reflecting low sample counts (Figure 5) and also the inaccuracy of the recording of some data fields in ECDS.

**Figure 5.**
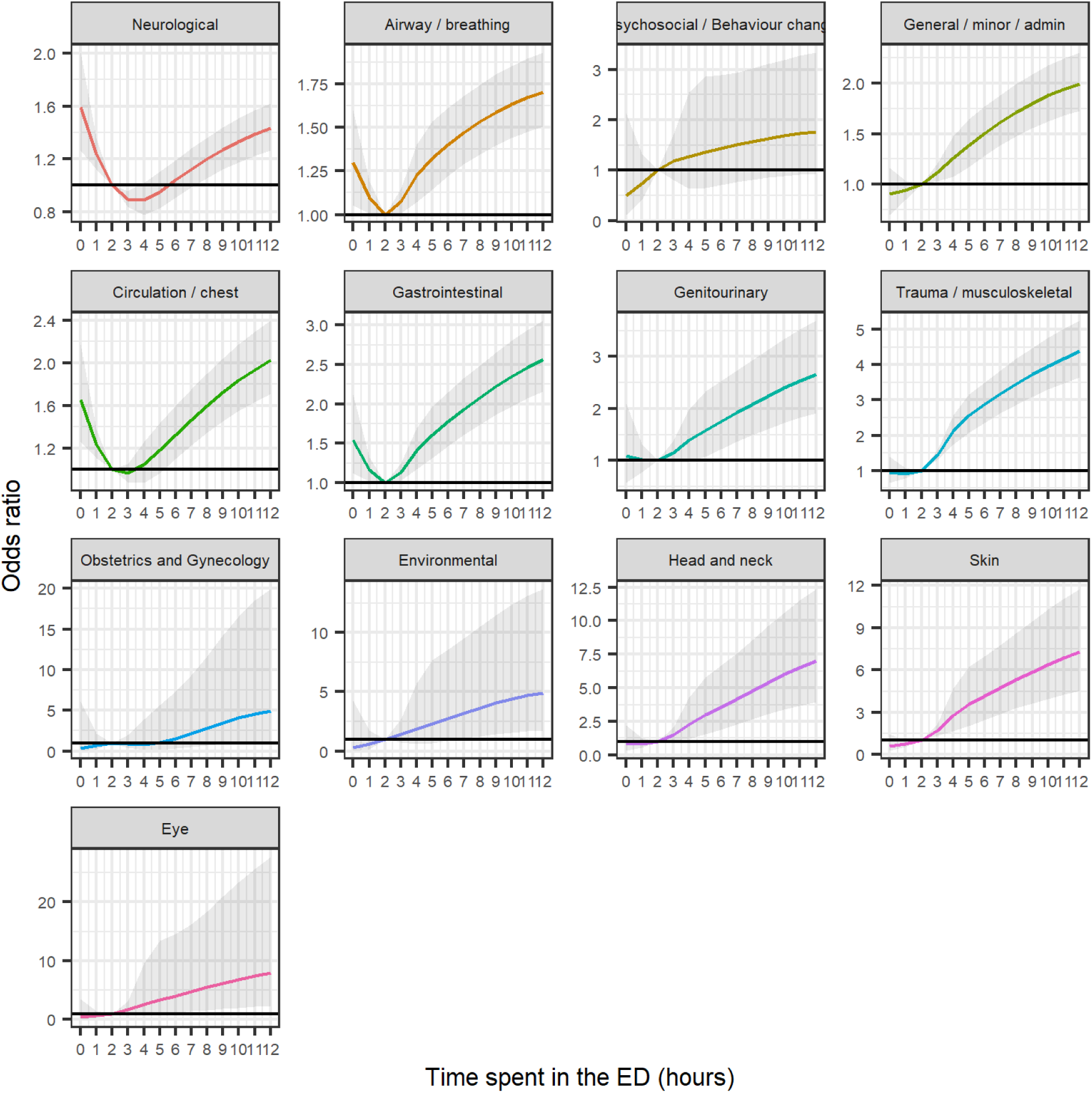
Odds ratios for all-cause, 30-day mortality as a function of time spent in the ED, by hospital chief complaint at arrival, compared with two hours Notes: 1. The estimates are adjusted for the covariates outlined in the Methods section. 2. The shaded area represents the 95% CI around the point estimates. 3. For presentation purposes, the y-axis range is not consistent between the panels due to differing effect sizes between groups.

**Figure 6.**
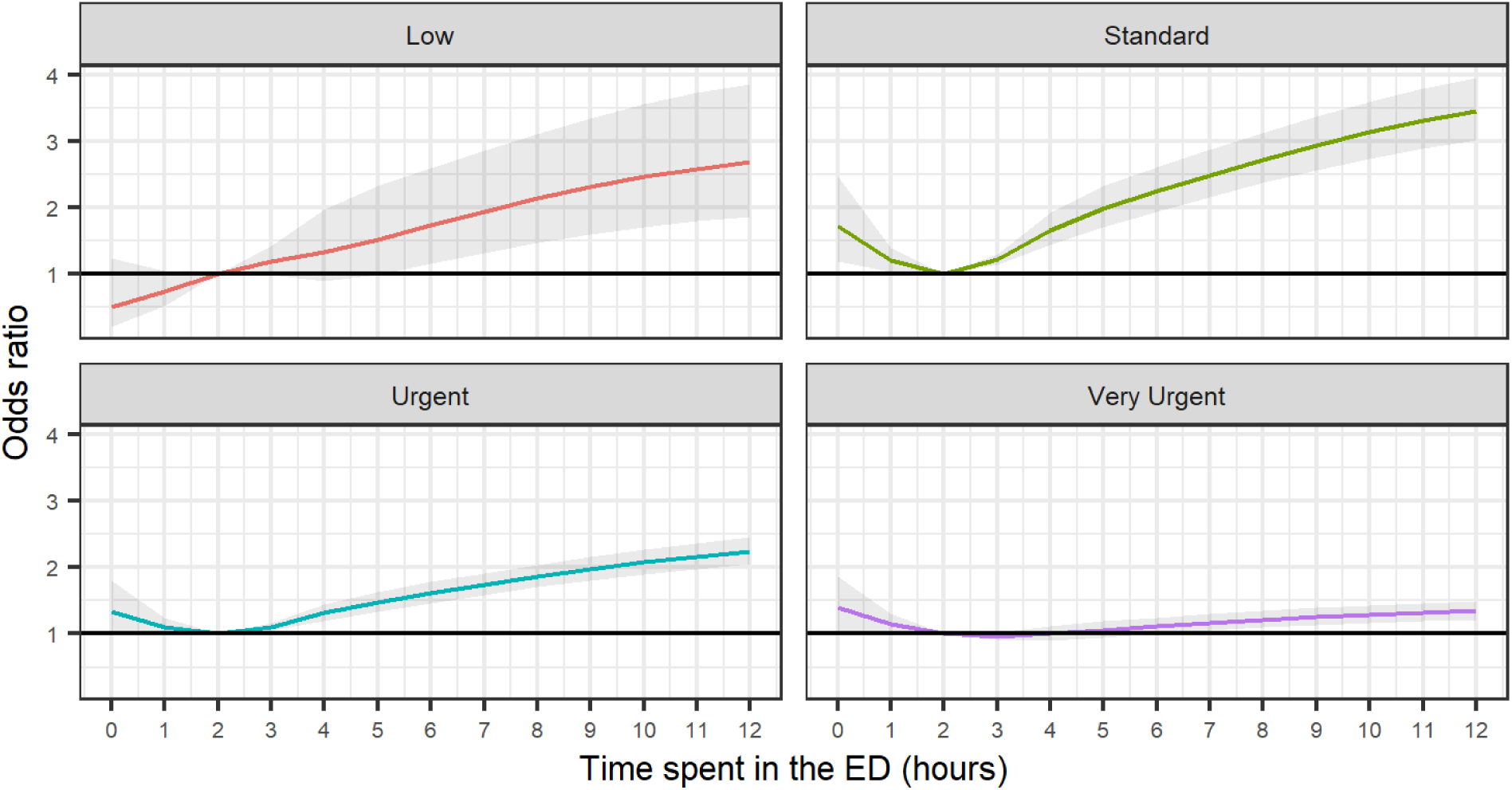
Odds ratios for all-cause, 30-day mortality as a function of time spent in the ED, by acuity group, compared with two hours Notes: 1. The estimates are adjusted for the covariates outlined in the Methods section. 2. The shaded area represents the 95% CI around the point estimates.

Individuals in the “standard” acuity group had the highest odds ratio for post-discharge mortality after 12 hours spent in an ED compared with two hours, with odds 3.4 (95% CI: 3.01 to 3.95) times higher. Individuals living in the “Very Urgent” acuity group had the smallest odds ratio for mortality after 12 hours compared to two hours, with odds 1.3 times (95% CI: 1.21 to 1.49) higher.

### Sensitivity analyses

Relative to the main model, including admission status as a covariate in the regression model decreased the adjusted odds ratios for post discharge death when compared with two hours to: 1.4 times higher for those who spent six hours in the ED (versus 1.6 times higher in the main model); 1.6 times higher for those who spent nine hours in the ED (versus 1.9 times higher in the main model); and 1.7 times higher for those who spent 12 hours in the ED (versus 2.1 times higher in the main model). See Supplementary Figure 9.

## Discussion

### Principal findings

We found that the risk of death within 30 days of leaving the ED (either to be discharged or admitted to inpatient hospital care) increased in a non-linear manner with total time spent in the ED for non-immediate treatment after approximately two hours. After 12 hours spent in the ED, the odds of post-discharge mortality were over twice as high compared to those spending two hours in the ED.

The relationship between time spent in the ED and the risk of post-discharge mortality was strongest for patient groups that generally experienced the lowest absolute risk of mortality: younger people, patients living in London, and patients that were not admitted to hospital.

### Research in context

Numerous studies have demonstrated the importance of timely emergency care. Our findings closely align with these studies, reaffirming the association between time spent in emergency care and the risk of mortality after leaving the department, not just during the visit. In Canada, the effect of overcrowding on extended waiting time in EDs has been shown to be associated with greater risk of short-term mortality or admission to hospital in lower acuity individuals [22]. In Australia, overcrowding has also been linked to increased mortality [23]. Similarly, in the UK it has been shown that delays to patient admission lasting more than five hours are associated with an increase in 30-day mortality, a trend that was demonstrated up to 12 hours, when data collection stopped because of unreliability [9].

The adverse effects of waiting time for healthcare services extend beyond emergency care settings. Delays in initiating cancer treatment have been shown to be associated with a significantly increased mortality rate, with every four-week delay increasing risk of death by 6-8% [24]. Poor health outcomes have also been demonstrated in smaller scale healthcare settings. Veterans that visited Veteran Affairs medical centres with facility-level waiting times over 31 days had significantly higher odds of mortality when compared to those waiting 31 days or fewer [25]. The impact of prolonged waits for healthcare also extends to mental health and social outcomes. Patients in the Netherlands waiting for elective general surgeries for varicose veins, inguinal hernia and gallstones were found to have increased anxiety, with 39-48% experiencing social life disruption and 18-23% facing work-related issues [26]. Negative emotional effects have also been shown for patients waiting for mental health services in UK [27], and in some cases, a deterioration of patient outcomes 12 months after treatment, with the strongest effects observed in people who waited for longer than three months [28].

Whilst our study results share similarities with those in the existing literature, there are differences in the methodologies used. For instance, our study looks at all patients whether they were discharged or admitted, rather than just the latter, and the total time spent in the ED, rather than just the time waiting for admission to an inpatient bed. This means that it covers a broader range of patients, not just the highest acuity patients on the admitted pathway. It also modelled time spent in the ED in a continuous rather than a discrete manner [9]; considered all deaths within 30-days after attendance; and had near-whole population coverage of England [22, 23].

### Strengths and limitations

Our research builds on previous studies demonstrating the association between time spent in an ED and post-discharge mortality by accounting for the patients’ comorbidities and chief complaint on arrival, which may otherwise skew the estimated relationship by introducing bias by confounding. We have also been able to show this trend on a considerably larger sample with wide population coverage, utilising current health and socio-economic information from the most recent national Census, meaning our results are estimated with considerable statistical precision and are likely to be generalisable to the broader population. By modelling time spent in the ED as a continuous variable, we avoided enforcing *a priori* groupings on the data, which may be difficult to justify and make it more difficult to identify inflexion points in the time-mortality relationship.

However, it is important to acknowledge several limitations of our study. We attempted to adjust for a wide range of confounding factors, both socio-demographic and health related, yet we cannot be sure that all confounding factors were completely adjusted for. For instance, we did not have access to a measure of ED overcrowding or changes in staffing, and whilst our socio-economic variables were derived from the latest available Census, this information could have become outdated for some characteristics and some groups by the end of the study period (for example, the highest qualification reported by school-age Census respondents). We were also unable to adjust for or assess the reasons for longer times spent in the ED; for example, some individuals may wait longer as they need to access specialist treatments, advice or services. In addition, we had no data at all on the physiological status of patients, such as their initial vital signs. Taken together, these considerations mean that the results of our study should be considered as estimates of statistical associations rather than necessarily cause-and-effect relationships.

To some extent, there is likely to be a lack of consistency and accuracy in the way patient data are captured in emergency care settings, particularly for recordings of sub-immediate acuity levels, which is ultimately a subjective clinical judgement. Whilst acuity is a reasonable proxy measure for condition severity, there is currently not a standardised way in which it is assessed and recorded, so it may vary from hospital to hospital and clinician to clinician. For this reason, we did not adjust for acuity level in our main model other than to remove the “immediate” acuity group and adjust for arrival by ambulance.

Our findings may not be generalisable to patients who genuinely spent more than 48 hours in an ED, those who had a missing acuity value on their record, and ED attendances beyond the first one for patients who visited an ED multiple times during the study period; these records were excluded from our analysis, which will hamper generalisability if they significantly differ from the included study population in terms of post-discharge mortality risk. We also excluded from the study population patients who required immediate care (resuscitation) upon arrival at the ED as residual confounding precluded reliable inferences being made, hence our findings are not necessarily generalisable to this group.

Finally, the study period for this analysis fell in the COVID-19 pandemic and overall, waiting times have worsened since then [29–30]. EDs implemented additional infection prevention control measures during the pandemic which may have affected the way that departments operated and may not be representative of how they operate today. However, ED attendance numbers had largely returned to their pre-pandemic level by June 2021 [31] after a fall the in earlier months of the pandemic. In addition, post-discharge mortality rates during the pandemic may be different to the current situation, for example because of relatively high levels of COVID-19-related mortality and social restrictions in place during the pandemic. The extent to which our findings are generalisable to the post-pandemic period is therefore uncertain.

### Conclusions

Our analysis builds on previous research to demonstrate that time spent in an ED may be an important risk factor for all-cause mortality within 30 days of being discharged or admitted to inpatient care, and not just during the ED attendance itself. These findings could be used by clinical decision-makers to inform strategies for improving patient flow through EDs, including improving triage processes and reducing “exit block” from the ED. These findings can also be used by policy makers when considering incentive systems and setting performance metrics for time spent in the ED. Future research is needed to understand the causal drivers of post-discharge mortality and to confirm that our findings are generalisable beyond the COVID-19 pandemic and in other urgent and emergency care settings.

## Data Availability

The source data are not publicly available and are subject to controlled access due to their sensitive nature. Census 2011 and death registration data are available through the Integrated Data Service (IDS). Details of the application requirements and process, and the use of data, are available at https://integrateddataservice.gov.uk/how-to-access-the-integrated-data-service. The NHS datasets that were used are held by NHS England and can be accessed through the NHS Secure Data Environment: https://digital.nhs.uk/services/secure-data-environment-service.

https://digital.nhs.uk/services/secure-data-environment-service

## References

[1] Office for National Statistics (ONS). Accident and Emergency wait times across the UK: 2024. 2024 https://www.ons.gov.uk/peoplepopulationandcommunity/healthandsocialcare/healthcaresystem/articles/accidentandemergencywaittimesacrosstheuk/2024-02-28

[2] Royal College of Emergency Medicine. RCEM acute insight series: crowding and its consequences. 2021. https://rcem.ac.uk/wp-content/uploads/2021/11/RCEM_Why_Emergency_Department_Crowding_Matters.pdf

[3] The King’s Fund. The number of hospital beds. 2023. https://www.kingsfund.org.uk/insight-and-analysis/data-and-charts/number-hospital-beds

[4] NHS England. NHS pressure continues as hospitals deal with high bed occupancy. 2023. https://www.england.nhs.uk/2023/01/nhs-pressure-continues-as-hospitals-deal-with-high-bed-occupancy/

[5] Paling S, Lambert J, Clouting J, González-Esquerré J, Auterson T. Waiting times in emergency departments: exploring the factors associated with longer patient waits for emergency care in England using routinely collected daily data. Emerg Med J. 2020 Dec;37(12):781–786. doi: 10.1136/emermed-2019-208849.

[6] Moskop JC, Sklar DP, Geiderman JM, Schears RM, Bookman KJ. Emergency department crowding, part 1-- concept, causes, and moral consequences. Ann Emerg Med. 2009 May;53(5):605–11. doi: 10.1016/j.annemergmed.2008.09.019.

[7] O’Dowd A. Growing pressure on NHS threatens frontline services. BMJ 2022; 379:o2439 doi:10.1136/bmj.o2439

[8] Institute for Fiscal Studies (IFS). IFS Green Budget 2021 Chapter 6-Pressures on the NHS. 2021. https://ifs.org.uk/sites/default/files/output_url_files/6-Pressures-on-the-NHS-.pdf

[9] Jones S, Moulton C, Swift S, Molyneux P, Black S, Mason N, Oakley R, Mann C. Association between delays to patient admission from the emergency department and all-cause 30-day mortality. Emerg Med J. 2022 Mar;39(3):168–173. doi: 10.1136/emermed-2021-211572.

[10] Plunkett PK, Byrne DG, Breslin T, Bennett K, Silke B. Increasing wait times predict increasing mortality for emergency medical admissions. Eur J Emerg Med. 2011 Aug;18(4):192–6. doi: 10.1097/MEJ.0b013e328344917e.

[11] Richardson DB. Increase in patient mortality at 10 days associated with emergency department overcrowding. Med J Aust. 2006;184(5):213–216. doi:10.5694/j.1326-5377.2006.tb00158.x.

[12] Singer AJ, Thode Jr HC, Viccellio P, Pines JM. The association between length of emergency department boarding and mortality. Acad Emerg Med. 2011;18(12):1324–1329. doi:10.1111/j.1553-2712.2011.01214.x.

[13] Office for National Statistics (ONS). Census 2021 to Personal Demographics Service (PDS) linkage report. 2023. https://www.ons.gov.uk/peoplepopulationandcommunity/healthandsocialcare/healthinequalities/methodologies/census2021topersonaldemographicsservicelinkagereport#:~:text=The%20linkage%20rate%20by%20census,of%20the%20census%20was%2096.93%25

[14] NHS Digital. Emergency Care Data Set (ECDS). https://digital.nhs.uk/data-and-information/data-collections-and-data-sets/data-sets/emergency-care-data-set-ecds

[15] NHS Digital. Hospital Episode Statistics. https://digital.nhs.uk/services/hospital-episode-statistics

[16] Office for National Statistics (ONS). Quality and methodology information for Census 2021. 2023. https://www.ons.gov.uk/peoplepopulationandcommunity/populationandmigration/populationestimates/methodologies/qualityandmethodologyinformationqmiforcensus2021

[17] Department for Communities and Local Government. English Index of Multiple Deprivation 2015: Guidance https://assets.publishing.service.gov.uk/media/5a7f0e5ded915d74e33f410b/English_Index_of_Multiple_Deprivation_2015_-_Guidance.pdf

[18] Office for National Statistics (ONS). The National Statistics Socio-economic Classification (NS-SEC) based on SOC2010. https://www.ons.gov.uk/methodology/classificationsandstandards/otherclassifications/thenationalstatisticssocioeconomicclassificationnssecrebasedonsoc2010

[19] Austin PC. An Introduction to Propensity Score Methods for Reducing the Effects of Confounding in Observational Studies. Multivariate Behav Res. 2011 May;46(3):399–424. doi: 10.1080/00273171.2011.568786.

[20] Perperoglou, A., Sauerbrei, W., Abrahamowicz, M. et al. A review of spline function procedures in R. BMC Med Res Methodol 19, 46 (2019). 10.1186/s12874-019-0666-3

[21] Russell V. Lenth. emmeans: Estimated Marginal Means, aka Least-Squares Means. R package version 1.8.5. 2023. https://CRAN.R-project.org/package=emmeans

[22] Guttmann A, Schull MJ, Vermeulen MJ, Stukel TA. Association between waiting times and short term mortality and hospital admission after departure from emergency department: population based cohort study from Ontario, Canada. BMJ. 2011;342:d2983. doi:10.1136/bmj.d2983.

[23] Sprivulis PC, Da Silva JA, Jacobs IG, Frazer AR, Jelinek GA. The association between hospital overcrowding and mortality among patients admitted via Western Australian emergency departments. Med J Aust. 2006 Mar 6;184(5):208–12. doi: 10.5694/j.1326-5377.2006.tb00416.x.

[24] Hanna T P, King W D, Thibodeau S, Jalink M, Paulin G A, Harvey-Jones E et al. Mortality due to cancer treatment delay: systematic review and meta-analysis. BMJ 2020;371:m4087 doi:10.1136/bmj.m4087

[25] Pizer SD, Prentice JC. What are the consequences of waiting for health care in the veteran population? J Gen Intern Med. 2011 Nov;26 Suppl 2:676–682. doi: 10.1007/s11606-011-1819-1.

[26] Oudhoff JP, Timmermans DR, Knol DL, Bijnen AB, van der Wal G. Waiting for elective general surgery: impact on health related quality of life and psychosocial consequences. BMC Public Health. 2007 Jul 19;7:164. doi: 10.1186/1471-2458-7-164.

[27] Punton G, Dodd AL, McNeill A. ’You’re on the waiting list’: An interpretive phenomenological analysis of young adults’ experiences of waiting lists within mental health services in the UK. PLoS One. 2022 Mar 18;17(3):e0265542. doi: 10.1371/journal.pone.0265542.

[28] Reichert A, Jacobs R. The impact of waiting time on patient outcomes: Evidence from early intervention in psychosis services in England. Health Econ. 2018 Nov;27(11):1772–1787. doi: 10.1002/hec.3800.

[29] Nuffield Trust. A&E waiting times. 2024. https://www.nuffieldtrust.org.uk/resource/a-e-waiting-times

[30] The King’s Fund. What’s going on with A&E waiting times? The King’s Fund. 2022. https://www.kingsfund.org.uk/publications/whats-going-ae-waiting-times

[31] RCEM Policy, 2024. https://public.flourish.studio/story/2813984/

