## Supplementary materials for "Association between time spent in the emergency department and 30-day mortality: a population-level observational study in England"

**Contents**

Supplementary Table 1. Variables used in modelling

Supplementary Table 2. Knot optimisation based on BIC

Supplementary Table 3. Number of patients that were alive or dead within 30 days of leaving the ED, by total time spent in the ED

Supplementary Table 4. Number of patients that were alive or dead within 30 days of leaving the ED, by total time spent in the ED and age group

Supplementary Table 5. Number of patients that were alive or dead within 30 days of leaving the ED, by total time spent in the ED and sex

Supplementary Table 6. Number of patients that were alive or dead within 30 days of leaving the ED, by total time spent in the ED and region of residence

Supplementary Table 7. Number of patients that were alive or dead within 30 days of leaving the ED, by total time spent in the ED and admission status

Supplementary Table 8. Number of patients that were alive or dead within 30 days of leaving the ED, by total time spent in the ED and relative area deprivation decile group

Supplementary Table 9. Number of patients that were alive or dead within 30 days of leaving the ED, by total time spent in the ED and chief complaint

Supplementary Table 10. Number of patients that were alive or dead within 30 days of leaving the ED, by total time spent in the ED and acuity level

Supplementary Figure 1. Estimated marginal probability of all-cause, 30-day mortality as a function of time spent in the ED, for individuals in the ‘immediate’ acuity group

Supplementary Figure 2. Estimated marginal probability of all-cause, 30-day mortality as a function of time to first treatment in the ED, for individuals in the ‘Very Urgent’, ‘Urgent’, ‘Standard’ and ‘Low’ acuity groups

Supplementary Figure 3. Sample flow diagram

Supplementary Figure 4. Distribution of total time spent in the ED up to 48 hours

Supplementary Figure 5. Adjusted odds ratios for all-cause, 30-day mortality as a function of time spent in the ED, compared to two hours

Supplementary Figure 6. Odds ratios compared to two hours for all-cause, 30-day mortality as a function of time spent in the ED, by age group, 12 hours

Supplementary Figure 7. Odds ratios compared to two hours for all-cause, 30-day mortality as a function of time spent in the ED, by region of residence, 12 hours

Supplementary Figure 8. Odds ratios compared to two hours for all-cause, 30-day mortality as a function of time spent in the ED, by relative area deprivation decile group, 48 hours

Supplementary Figure 9. Odds ratios compared to two hours for all-cause, 30-day mortality as a function of time spent in the ED, adjusted for admission status – sensitivity analysis including admission status as a covariate in the model

**Supplementary Table 1. Variables used in modelling**

| **Variable** | **Coding** | **Source** |
| --- | --- | --- |
| **Age** | Single year of age | ECDS |
| **Arrival month** | January, February, March, April, May, June, July, August, September, October, November, December | ECDS |
| **Arrival time of day** | 12am to 6am, 6am to 12pm, 12pm to 6pm, 6pm to 12am | ECDS |
| **Sex** | Male, female | Census 2021 |
| **Ethnicity** | White, Asian, Black, Mixed, Other | Census 2021 |
| **Region** | East Midlands, East of England, London, North East England, North West England, South East England, South West England, West Midlands, Yorkshire and The Humber | Census 2021 |
| **Local area deprivation decile group** | Deciles of deprivation:  1 (most deprived), 2, 3, 4, 5, 6, 7, 8, 9, 10 (least deprived | Census 2021 |
| **National Statistics Socio-Economic Classification** | Higher managerial, Lower managerial, Intermediate occupations, Smaller employers, Lower supervisory, Semi-routine, Routine occupations, Never worked, No code required | Census 2021 |
| **Highest qualification** | No qualifications, 1-4 GCSEs/O-levels, 5+ GCSEs/O-levels, apprenticeship, 2+ A-levels or equivalent, degree or above, other qualification, No code required | Census 2021 |
| **Self-reported health** | Very good, Good, Fair, Bad, Very bad | Census 2021 |
| **Self-reported long-term health condition or disability** | No long-term health conditions, Day-to-day activities not reduced at all by long-term health conditions, Day-to-day activities reduced a little by long-term health conditions, Day-to-day activities reduced a lot by long-term health conditions | Census 2021 |
| **Chief complaint** | General / minor / admin, Airway / breathing, Circulation / chest, Environmental, Eye, Gastrointestinal, Genitourinary, Head and neck, Neurological, Obstetrics and gynaecology, Psychosocial / Behaviour change, Skin, Trauma / musculoskeletal | ECDS |
| **Mode of arrival** | Emergency ambulance, Not emergency ambulance | ECDS |
| **Admission to hospital status** | Admitted, Not admitted | ECDS |
| **Comorbidity: Covid** | Ever, Never | HES APC / OP |
| **Comorbidity: Cancer** | Ever, Never | HES APC / OP |
| **Comorbidity: Diabetes** | Ever, Never | HES APC / OP |
| **Comorbidity: Dementia** | Ever, Never | HES APC / OP |
| **Comorbidity: Serious mental illness** | Ever, Never | HES APC / OP |
| **Comorbidity: Autism** | Ever, Never | HES APC / OP |
| **Comorbidity: Neurological MND, Parkinson’s MS** | Ever, Never | HES APC / OP |
| **Comorbidity: Alzheimer** | Ever, Never | HES APC / OP |
| **Comorbidity: Epilepsy** | Ever, Never | HES APC / OP |
| **Comorbidity: Hypertension** | Ever, Never | HES APC / OP |
| **Comorbidity: Angina** | Ever, Never | HES APC / OP |
| **Comorbidity: Myocardial Infarction** | Ever, Never | HES APC / OP |
| **Comorbidity: Ischaemic hear disease** | Ever, Never | HES APC / OP |
| **Comorbidity: Atrial fibrillation** | Ever, Never | HES APC / OP |
| **Comorbidity: Heart failure** | Ever, Never | HES APC / OP |
| **Comorbidity: Stroke** | Ever, Never | HES APC / OP |
| **Comorbidity: Other respiratory infection** | Ever, Never | HES APC / OP |
| **Comorbidity: Influenza pneumonia** | Ever, Never | HES APC / OP |
| **Comorbidity: Chronic obstructive pulmonary disease and respiratory failure** | Ever, Never | HES APC / OP |
| **Comorbidity: Asthma** | Ever, Never | HES APC / OP |
| **Comorbidity: Inflammatory bowel disease** | Ever, Never | HES APC / OP |
| **Comorbidity: Liver disease** | Ever, Never | HES APC / OP |
| **Comorbidity: Rheumatoid arthritis** | Ever, Never | HES APC / OP |
| **Comorbidity: Osteoarthritis** | Ever, Never | HES APC / OP |
| **Comorbidity: Osteoporosis** | Ever, Never | HES APC / OP |
| **Comorbidity: Kidney disease** | Ever, Never | HES APC / OP |

ECDS: Emergency Care Dataset; HES APC / OP: Hospital Episode Statistics Admitted Patient Care / Outpatient.

**Supplementary** **Table 2. Knot optimisation based on BIC**

| **Model term** | **Position of knots (percentiles)** | **BIC score** |
| --- | --- | --- |
| **Age: internal knots** | 0.50 | 683639.25 |
|  | 0.33, 0.67 | 683647.14 |
|  | 0.25, 0.50, 0.75 | 683556.97 |
|  | 0.20, 0.40, 0.60, 0.80 | 683517.90 |
|  | 0.10, 0.25, 0.50, 0.75, 0.90 | 683493.34 |
| **Age: boundary knots** | 0.00, 1.00 | 683498.64 |
|  | 0.01, 0.99 | 683517.73 |
|  | 0.05, 0.95 | 683549.71 |
|  | 0.10, 0.90 | 683574.22 |
| **Time spent in the ED: internal knots** | 0.50 | 683564.13 |
|  | 0.33, 0.67 | 683552.86 |
|  | 0.25, 0.50, 0.75 | 683498.64 |
|  | 0.20, 0.40, 0.60, 0.80 | 683515.57 |
|  | 0.10, 0.25, 0.50, 0.75, 0.90 | 683523.23 |
| **Time spent in the ED: boundary knots** | 0.00, 1.00 | 683663.00 |
|  | 0.01, 0.99 | 683582.68 |
|  | 0.05, 0.95 | 683613.33 |
|  | 0.10, 0.90 | 683730.67 |

BIC: Bayesian Information Criterion

**Supplementary Table 3. Number of patients that were alive or dead within 30 days of leaving the ED by total time spent in the ED**

| **Total time spent in the ED (hours, rounded to the nearest hour)** | **Alive or dead within 30 days of discharge** | **Sample Size** |
| --- | --- | --- |
| **0hr to 59m** | Alive | 156,430 |
|  | Dead | 500 |
| **1** | Alive | 714,366 |
|  | Dead | 1,255 |
| **2** | Alive | 1,092,765 |
|  | Dead | 2,959 |
| **3** | Alive | 1,281,272 |
|  | Dead | 6,912 |
| **4** | Alive | 1,247,259 |
|  | Dead | 13,330 |
| **5** | Alive | 516,091 |
|  | Dead | 7,767 |
| **6** | Alive | 420,983 |
|  | Dead | 8,228 |
| **7** | Alive | 298,121 |
|  | Dead | 7,246 |
| **8** | Alive | 218,492 |
|  | Dead | 6,503 |
| **9** | Alive | 154,588 |
|  | Dead | 5,377 |
| **10** | Alive | 115,558 |
|  | Dead | 4,574 |
| **11** | Alive | 84,059 |
|  | Dead | 3,852 |
| **12** | Alive | 65,357 |
|  | Dead | 3,447 |
| **13 or more** | Alive | 267,181 |
|  | Dead | 16,707 |

**Supplementary Table 4. Number of patients that were alive or dead within 30 days of leaving the ED, by total time spent in the ED and age group**

| **Total time spent in the ED (hours, rounded to the nearest hour)** | **Age group** | **Alive or dead within 30 days of discharge** | **Sample Size** |
| --- | --- | --- | --- |
| **<2** | <20 years old | Alive | 351,020 |
|  |  | Dead | 18 |
|  | 20 to 39 years old | Alive | 228,539 |
|  |  | Dead | 32 |
|  | 40 to 59 years old | Alive | 171,471 |
|  |  | Dead | 192 |
|  | 60 to 79 years old | Alive | 97,181 |
|  |  | Dead | 707 |
|  | ≥80 years old | Alive | 22,585 |
|  |  | Dead | 806 |
| **2 and 3** | <20 years old | Alive | 876,491 |
|  |  | Dead | 49 |
|  | 20 to 39 years old | Alive | 574,547 |
|  |  | Dead | 173 |
|  | 40 to 59 years old | Alive | 477,047 |
|  |  | Dead | 870 |
|  | 60 to 79 years old | Alive | 331,870 |
|  |  | Dead | 3,759 |
|  | ≥80 years old | Alive | 114,082 |
|  |  | Dead | 5,020 |
| **4 and 5** | <20 years old | Alive | 422,200 |
|  |  | Dead | 48 |
|  | 20 to 39 years old | Alive | 409,864 |
|  |  | Dead | 203 |
|  | 40 to 59 years old | Alive | 390,226 |
|  |  | Dead | 1,576 |
|  | 60 to 79 years old | Alive | 360,063 |
|  |  | Dead | 7,813 |
|  | ≥80 years old | Alive | 180,997 |
|  |  | Dead | 11,457 |
| **6 and 7** | <20 | Alive | 109,943 |
|  |  | Dead | 26 |
|  | 20 to 39 years old | Alive | 155,180 |
|  |  | Dead | 124 |
|  | 40 to 59 years old | Alive | 164,818 |
|  |  | Dead | 1,050 |
|  | 60 to 79 years old | Alive | 181,859 |
|  |  | Dead | 5,724 |
|  | ≥80 years old | Alive | 107,304 |
|  |  | Dead | 8,550 |
| **8 to 11** | <20 years old | Alive | 54,437 |
|  |  | Dead | 12 |
|  | 20 to 39 years old | Alive | 106,362 |
|  |  | Dead | 164 |
|  | 40 to 59 years old | Alive | 129,306 |
|  |  | Dead | 1,378 |
|  | 60 to 79 years old | Alive | 169,786 |
|  |  | Dead | 7,425 |
|  | ≥80 years old | Alive | 112,806 |
|  |  | Dead | 11,327 |
| **12 to 48** | <20 years old | Alive | 15,461 |
|  |  | Dead | 10 |
|  | 20 to 39 years old | Alive | 42,198 |
|  |  | Dead | 139 |
|  | 40 to 59 years old | Alive | 65,797 |
|  |  | Dead | 1,196 |
|  | 60 to 79 years old | Alive | 117,013 |
|  |  | Dead | 7,424 |
|  | ≥80 years old | Alive | 92,069 |
|  |  | Dead | 11,385 |

Notes:

Total time spent in the ED has been aggregated into groups due to low counts.

**Supplementary Table 5. Number of patients that were alive or dead within 30 days of leaving the ED, by total time spent in the ED and sex**

| **Total time spent in the ED (hours, rounded to the nearest hour)** | **Sex** | **Alive or dead within 30 days of discharge** | **Sample Size** |
| --- | --- | --- | --- |
| **0hr to 59m** | Male | Alive | 74,378 |
|  |  | Dead | 262 |
|  | Female | Alive | 82,052 |
|  |  | Dead | 238 |
| **1** | Male | Alive | 360,366 |
|  |  | Dead | 678 |
|  | Female | Alive | 354,000 |
|  |  | Dead | 577 |
| **2** | Male | Alive | 548,856 |
|  |  | Dead | 1,578 |
|  | Female | Alive | 543,909 |
|  |  | Dead | 1,381 |
| **3** | Male | Alive | 615,710 |
|  |  | Dead | 3,659 |
|  | Female | Alive | 665,562 |
|  |  | Dead | 3,253 |
| **4** | Male | Alive | 574,513 |
|  |  | Dead | 6,945 |
|  | Female | Alive | 672,746 |
|  |  | Dead | 6,385 |
| **5** | Male | Alive | 234,076 |
|  |  | Dead | 4,132 |
|  | Female | Alive | 282,015 |
|  |  | Dead | 3,635 |
| **6** | Male | Alive | 189,037 |
|  |  | Dead | 4,198 |
|  | Female | Alive | 231,946 |
|  |  | Dead | 4,030 |
| **7** | Male | Alive | 133,532 |
|  |  | Dead | 3,686 |
|  | Female | Alive | 164,589 |
|  |  | Dead | 3,560 |
| **8** | Male | Alive | 97,540 |
|  |  | Dead | 3,349 |
|  | Female | Alive | 120,952 |
|  |  | Dead | 3,154 |
| **9** | Male | Alive | 69,570 |
|  |  | Dead | 2,698 |
|  | Female | Alive | 85,018 |
|  |  | Dead | 2,679 |
| **10** | Male | Alive | 51,781 |
|  |  | Dead | 2,392 |
|  | Female | Alive | 63,777 |
|  |  | Dead | 2,182 |
| **11** | Male | Alive | 37,979 |
|  |  | Dead | 2,040 |
|  | Female | Alive | 46,080 |
|  |  | Dead | 1,812 |
| **12** | Male | Alive | 29,574 |
|  |  | Dead | 1,732 |
|  | Female | Alive | 35,783 |
|  |  | Dead | 1,715 |
| **13 to 48** | Male | Alive | 120,383 |
|  |  | Dead | 8,490 |
|  | Female | Alive | 146,798 |
|  |  | Dead | 8,217 |

Notes:

Total time spent in the ED >12 hours has been aggregated due to low counts.

**Supplementary Table 6. Number of patients that were alive or dead within 30 days of leaving the ED, by total time spent in the ED and region of residence**

| **Total time spent in the ED (hours, rounded to the nearest hour)** | **Region of residence** | **Alive or dead within 30 days of discharge** | **Sample Size** |
| --- | --- | --- | --- |
| **0hr to 59m** | East Midlands | Alive | 16,978 |
|  |  | Dead | 31 |
|  | East of England | Alive | 10,056 |
|  |  | Dead | 23 |
|  | London | Alive | 15,048 |
|  |  | Dead | 16 |
|  | North East | Alive | 19,476 |
|  |  | Dead | 35 |
|  | North West | Alive | 18,831 |
|  |  | Dead | 43 |
|  | South East | Alive | 13,264 |
|  |  | Dead | 36 |
|  | South West | Alive | 10,342 |
|  |  | Dead | 34 |
|  | West Midlands | Alive | 20,468 |
|  |  | Dead | 238 |
|  | Yorkshire and The Humber | Alive | 31,967 |
|  |  | Dead | 44 |
| **1** | East Midlands | Alive | 76,895 |
|  |  | Dead | 150 |
|  | East of England | Alive | 78,708 |
|  |  | Dead | 130 |
|  | London | Alive | 65,647 |
|  |  | Dead | 60 |
|  | North East | Alive | 35,164 |
|  |  | Dead | 84 |
|  | North West | Alive | 99,895 |
|  |  | Dead | 173 |
|  | South East | Alive | 82,560 |
|  |  | Dead | 145 |
|  | South West | Alive | 56,812 |
|  |  | Dead | 166 |
|  | West Midlands | Alive | 99,664 |
|  |  | Dead | 193 |
|  | Yorkshire and The Humber | Alive | 119,021 |
|  |  | Dead | 154 |
| **2** | East Midlands | Alive | 99,796 |
|  |  | Dead | 273 |
|  | East of England | Alive | 132,988 |
|  |  | Dead | 333 |
|  | London | Alive | 125,282 |
|  |  | Dead | 152 |
|  | North East | Alive | 48,153 |
|  |  | Dead | 224 |
|  | North West | Alive | 153,866 |
|  |  | Dead | 398 |
|  | South East | Alive | 141,500 |
|  |  | Dead | 456 |
|  | South West | Alive | 96,305 |
|  |  | Dead | 409 |
|  | West Midlands | Alive | 145,677 |
|  |  | Dead | 354 |
|  | Yorkshire and The Humber | Alive | 149,198 |
|  |  | Dead | 360 |
| **3** | East Midlands | Alive | 107,356 |
|  |  | Dead | 666 |
|  | East of England | Alive | 153,029 |
|  |  | Dead | 807 |
|  | London | Alive | 184,556 |
|  |  | Dead | 473 |
|  | North East | Alive | 62,695 |
|  |  | Dead | 712 |
|  | North West | Alive | 170,646 |
|  |  | Dead | 789 |
|  | South East | Alive | 182,305 |
|  |  | Dead | 1,104 |
|  | South West | Alive | 115,767 |
|  |  | Dead | 889 |
|  | West Midlands | Alive | 156,523 |
|  |  | Dead | 725 |
|  | Yorkshire and The Humber | Alive | 148,395 |
|  |  | Dead | 747 |
| **4** | East Midlands | Alive | 96,433 |
|  |  | Dead | 1,342 |
|  | East of England | Alive | 138,061 |
|  |  | Dead | 1,643 |
|  | London | Alive | 208,684 |
|  |  | Dead | 1,133 |
|  | North East | Alive | 69,079 |
|  |  | Dead | 1,620 |
|  | North West | Alive | 157,268 |
|  |  | Dead | 1,398 |
|  | South East | Alive | 198,896 |
|  |  | Dead | 2,141 |
|  | South West | Alive | 111,896 |
|  |  | Dead | 1,480 |
|  | West Midlands | Alive | 130,876 |
|  |  | Dead | 1,238 |
|  | Yorkshire and The Humber | Alive | 136,066 |
|  |  | Dead | 1,335 |
| **5** | East Midlands | Alive | 48,608 |
|  |  | Dead | 809 |
|  | East of England | Alive | 61,160 |
|  |  | Dead | 963 |
|  | London | Alive | 61,427 |
|  |  | Dead | 380 |
|  | North East | Alive | 23,843 |
|  |  | Dead | 624 |
|  | North West | Alive | 67,548 |
|  |  | Dead | 880 |
|  | South East | Alive | 74,551 |
|  |  | Dead | 1,068 |
|  | South West | Alive | 54,454 |
|  |  | Dead | 1,043 |
|  | West Midlands | Alive | 64,365 |
|  |  | Dead | 983 |
|  | Yorkshire and The Humber | Alive | 60,135 |
|  |  | Dead | 1,017 |
| **6** | East Midlands | Alive | 39,078 |
|  |  | Dead | 848 |
|  | East of England | Alive | 48,218 |
|  |  | Dead | 1,014 |
|  | London | Alive | 53,347 |
|  |  | Dead | 509 |
|  | North East | Alive | 19,004 |
|  |  | Dead | 627 |
|  | North West | Alive | 57,191 |
|  |  | Dead | 1,048 |
|  | South East | Alive | 62,603 |
|  |  | Dead | 1,110 |
|  | South West | Alive | 42,797 |
|  |  | Dead | 1,024 |
|  | West Midlands | Alive | 50,990 |
|  |  | Dead | 990 |
|  | Yorkshire and The Humber | Alive | 47,755 |
|  |  | Dead | 1,058 |
| **7** | East Midlands | Alive | 29,160 |
|  |  | Dead | 790 |
|  | East of England | Alive | 34,098 |
|  |  | Dead | 895 |
|  | London | Alive | 39,290 |
|  |  | Dead | 505 |
|  | North East | Alive | 12,335 |
|  |  | Dead | 455 |
|  | North West | Alive | 41,789 |
|  |  | Dead | 975 |
|  | South East | Alive | 41,610 |
|  |  | Dead | 961 |
|  | South West | Alive | 29,862 |
|  |  | Dead | 829 |
|  | West Midlands | Alive | 36,291 |
|  |  | Dead | 889 |
|  | Yorkshire and The Humber | Alive | 33,686 |
|  |  | Dead | 947 |
| **8** | East Midlands | Alive | 22,331 |
|  |  | Dead | 654 |
|  | East of England | Alive | 24,681 |
|  |  | Dead | 725 |
|  | London | Alive | 28,771 |
|  |  | Dead | 492 |
|  | North East | Alive | 8,561 |
|  |  | Dead | 431 |
|  | North West | Alive | 32,033 |
|  |  | Dead | 921 |
|  | South East | Alive | 29,537 |
|  |  | Dead | 847 |
|  | South West | Alive | 21,593 |
|  |  | Dead | 736 |
|  | West Midlands | Alive | 26,607 |
|  |  | Dead | 811 |
|  | Yorkshire and The Humber | Alive | 24,378 |
|  |  | Dead | 886 |
| **9** | East Midlands | Alive | 16,536 |
|  |  | Dead | 596 |
|  | East of England | Alive | 17,331 |
|  |  | Dead | 608 |
|  | London | Alive | 20,604 |
|  |  | Dead | 383 |
|  | North East | Alive | 5,397 |
|  |  | Dead | 287 |
|  | North West | Alive | 23,507 |
|  |  | Dead | 804 |
|  | South East | Alive | 19,984 |
|  |  | Dead | 644 |
|  | South West | Alive | 14,881 |
|  |  | Dead | 628 |
|  | West Midlands | Alive | 19,160 |
|  |  | Dead | 711 |
|  | Yorkshire and The Humber | Alive | 17,188 |
|  |  | Dead | 716 |
| **10** | East Midlands | Alive | 12,567 |
|  |  | Dead | 496 |
|  | East of England | Alive | 12,582 |
|  |  | Dead | 509 |
|  | London | Alive | 15,216 |
|  |  | Dead | 319 |
|  | North East | Alive | 3,951 |
|  |  | Dead | 214 |
|  | North West | Alive | 18,565 |
|  |  | Dead | 736 |
|  | South East | Alive | 14,690 |
|  |  | Dead | 606 |
|  | South West | Alive | 11,073 |
|  |  | Dead | 505 |
|  | West Midlands | Alive | 14,480 |
|  |  | Dead | 597 |
|  | Yorkshire and The Humber | Alive | 12,434 |
|  |  | Dead | 592 |
| **11** | East Midlands | Alive | 9,509 |
|  |  | Dead | 463 |
|  | East of England | Alive | 9,151 |
|  |  | Dead | 384 |
|  | London | Alive | 10,816 |
|  |  | Dead | 265 |
|  | North East | Alive | 2,685 |
|  |  | Dead | 165 |
|  | North West | Alive | 13,987 |
|  |  | Dead | 637 |
|  | South East | Alive | 10,401 |
|  |  | Dead | 484 |
|  | South West | Alive | 7,926 |
|  |  | Dead | 404 |
|  | West Midlands | Alive | 10,611 |
|  |  | Dead | 550 |
|  | Yorkshire and The Humber | Alive | 8,973 |
|  |  | Dead | 500 |
| **12** | East Midlands | Alive | 7,559 |
|  |  | Dead | 412 |
|  | East of England | Alive | 7,037 |
|  |  | Dead | 393 |
|  | London | Alive | 8,064 |
|  |  | Dead | 251 |
|  | North East | Alive | 1,901 |
|  |  | Dead | 140 |
|  | North West | Alive | 11,424 |
|  |  | Dead | 614 |
|  | South East | Alive | 7,876 |
|  |  | Dead | 390 |
|  | South West | Alive | 6,215 |
|  |  | Dead | 346 |
|  | West Midlands | Alive | 8,308 |
|  |  | Dead | 457 |
|  | Yorkshire and The Humber | Alive | 6,973 |
|  |  | Dead | 444 |
| **13 to 48** | East Midlands | Alive | 29,053 |
|  |  | Dead | 2,011 |
|  | East of England | Alive | 27,615 |
|  |  | Dead | 1,498 |
|  | London | Alive | 35,111 |
|  |  | Dead | 1,311 |
|  | North East | Alive | 3,803 |
|  |  | Dead | 322 |
|  | North West | Alive | 55,756 |
|  |  | Dead | 3,791 |
|  | South East | Alive | 32,849 |
|  |  | Dead | 1,922 |
|  | South West | Alive | 26,955 |
|  |  | Dead | 2,040 |
|  | West Midlands | Alive | 34,767 |
|  |  | Dead | 2,353 |
|  | Yorkshire and The Humber | Alive | 21,272 |
|  |  | Dead | 1,459 |

Notes:

Total time spent in the ED >12 hours has been aggregated due to low counts.

**Supplementary Table 7. Number of patients that were alive or dead within 30 days of leaving the ED, by total time spent in the ED and admission status**

| **Total time spent in the ED (hours, rounded to the nearest hour)** | **Admission status** | **Alive or dead within 30 days of discharge** | **Sample Size** |
| --- | --- | --- | --- |
| **0hr to 59m** | Admitted | Alive | 50,637 |
|  |  | Dead | 360 |
|  | Not Admitted | Alive | 105,793 |
|  |  | Dead | 140 |
| **1** | Admitted | Alive | 108,784 |
|  |  | Dead | 862 |
|  | Not Admitted | Alive | 605,582 |
|  |  | Dead | 393 |
| **2** | Admitted | Alive | 111,438 |
|  |  | Dead | 2,090 |
|  | Not Admitted | Alive | 981,327 |
|  |  | Dead | 869 |
| **3** | Admitted | Alive | 175,780 |
|  |  | Dead | 5,175 |
|  | Not Admitted | Alive | 1,105,492 |
|  |  | Dead | 1,737 |
| **4** | Admitted | Alive | 296,349 |
|  |  | Dead | 10,572 |
|  | Not Admitted | Alive | 950,910 |
|  |  | Dead | 2,758 |
| **5** | Admitted | Alive | 133,127 |
|  |  | Dead | 6,073 |
|  | Not Admitted | Alive | 382,964 |
|  |  | Dead | 1,694 |
| **6** | Admitted | Alive | 132,378 |
|  |  | Dead | 6,592 |
|  | Not Admitted | Alive | 288,605 |
|  |  | Dead | 1,636 |
| **7** | Admitted | Alive | 110,647 |
|  |  | Dead | 5,869 |
|  | Not Admitted | Alive | 187,474 |
|  |  | Dead | 1,377 |
| **8** | Admitted | Alive | 94,217 |
|  |  | Dead | 5,405 |
|  | Not Admitted | Alive | 124,275 |
|  |  | Dead | 1,098 |
| **9** | Admitted | Alive | 75,500 |
|  |  | Dead | 4,561 |
|  | Not Admitted | Alive | 79,088 |
|  |  | Dead | 816 |
| **10** | Admitted | Alive | 62,546 |
|  |  | Dead | 3,922 |
|  | Not Admitted | Alive | 53,012 |
|  |  | Dead | 652 |
| **11** | Admitted | Alive | 49,932 |
|  |  | Dead | 3,332 |
|  | Not Admitted | Alive | 34,127 |
|  |  | Dead | 520 |
| **12** | Admitted | Alive | 42,276 |
|  |  | Dead | 2,998 |
|  | Not Admitted | Alive | 23,081 |
|  |  | Dead | 449 |
| **13 to 48** | Admitted | Alive | 195,933 |
|  |  | Dead | 14,757 |
|  | Not Admitted | Alive | 71,248 |
|  |  | Dead | 1,950 |

Notes:

Total time spent in the ED >12 hours has been aggregated due to low counts.

**Supplementary Table 8. Number of patients that were alive or dead within 30 days of leaving the ED, by total time spent in the ED and relative area deprivation decile group**

| **Total time spent in the ED (hours, rounded to the nearest hour)** | **Area deprivation decile group** | **Alive or dead within 30 days of discharge** | **Sample Size** |
| --- | --- | --- | --- |
| **0hr to 59m** | 1 (most deprived) | Alive | 26,902 |
|  |  | Dead | 51 |
|  | 2 | Alive | 20,564 |
|  |  | Dead | 65 |
|  | 3 | Alive | 17,617 |
|  |  | Dead | 54 |
|  | 4 | Alive | 15,278 |
|  |  | Dead | 51 |
|  | 5 | Alive | 14,252 |
|  |  | Dead | 42 |
|  | 6 | Alive | 13,821 |
|  |  | Dead | 47 |
|  | 7 | Alive | 13,312 |
|  |  | Dead | 46 |
|  | 8 | Alive | 12,528 |
|  |  | Dead | 46 |
|  | 9 | Alive | 11,632 |
|  |  | Dead | 42 |
|  | 10 (least deprived) | Alive | 10,524 |
|  |  | Dead | 56 |
| **1** | 1 (most deprived) | Alive | 98,501 |
|  |  | Dead | 119 |
|  | 2 | Alive | 84,776 |
|  |  | Dead | 116 |
|  | 3 | Alive | 77,410 |
|  |  | Dead | 99 |
|  | 4 | Alive | 70,970 |
|  |  | Dead | 121 |
|  | 5 | Alive | 69,234 |
|  |  | Dead | 143 |
|  | 6 | Alive | 67,887 |
|  |  | Dead | 148 |
|  | 7 | Alive | 63,693 |
|  |  | Dead | 124 |
|  | 8 | Alive | 64,124 |
|  |  | Dead | 129 |
|  | 9 | Alive | 60,108 |
|  |  | Dead | 127 |
|  | 10 (least deprived) | Alive | 57,663 |
|  |  | Dead | 129 |
| **2** | 1 (most deprived) | Alive | 141,749 |
|  |  | Dead | 280 |
|  | 2 | Alive | 126,508 |
|  |  | Dead | 265 |
|  | 3 | Alive | 119,706 |
|  |  | Dead | 262 |
|  | 4 | Alive | 110,564 |
|  |  | Dead | 304 |
|  | 5 | Alive | 107,896 |
|  |  | Dead | 325 |
|  | 6 | Alive | 104,110 |
|  |  | Dead | 323 |
|  | 7 | Alive | 99,553 |
|  |  | Dead | 293 |
|  | 8 | Alive | 99,013 |
|  |  | Dead | 323 |
|  | 9 | Alive | 93,988 |
|  |  | Dead | 292 |
|  | 10 (least deprived) | Alive | 89,678 |
|  |  | Dead | 292 |
| **3** | 1 (most deprived) | Alive | 157,181 |
|  |  | Dead | 666 |
|  | 2 | Alive | 148,032 |
|  |  | Dead | 635 |
|  | 3 | Alive | 143,917 |
|  |  | Dead | 629 |
|  | 4 | Alive | 134,029 |
|  |  | Dead | 714 |
|  | 5 | Alive | 127,953 |
|  |  | Dead | 751 |
|  | 6 | Alive | 124,365 |
|  |  | Dead | 739 |
|  | 7 | Alive | 117,520 |
|  |  | Dead | 711 |
|  | 8 | Alive | 114,498 |
|  |  | Dead | 733 |
|  | 9 | Alive | 110,095 |
|  |  | Dead | 686 |
|  | 10 (least deprived) | Alive | 103,682 |
|  |  | Dead | 648 |
| **4** | 1 (most deprived) | Alive | 146,622 |
|  |  | Dead | 1,152 |
|  | 2 | Alive | 141,434 |
|  |  | Dead | 1,247 |
|  | 3 | Alive | 139,607 |
|  |  | Dead | 1,367 |
|  | 4 | Alive | 132,734 |
|  |  | Dead | 1,376 |
|  | 5 | Alive | 125,955 |
|  |  | Dead | 1,417 |
|  | 6 | Alive | 122,456 |
|  |  | Dead | 1,489 |
|  | 7 | Alive | 114,782 |
|  |  | Dead | 1,325 |
|  | 8 | Alive | 111,948 |
|  |  | Dead | 1,364 |
|  | 9 | Alive | 108,808 |
|  |  | Dead | 1,390 |
|  | 10 (least deprived) | Alive | 102,913 |
|  |  | Dead | 1,203 |
| **5** | 1 (most deprived) | Alive | 63,210 |
|  |  | Dead | 761 |
|  | 2 | Alive | 57,290 |
|  |  | Dead | 810 |
|  | 3 | Alive | 54,404 |
|  |  | Dead | 712 |
|  | 4 | Alive | 53,098 |
|  |  | Dead | 774 |
|  | 5 | Alive | 50,825 |
|  |  | Dead | 810 |
|  | 6 | Alive | 50,301 |
|  |  | Dead | 792 |
|  | 7 | Alive | 48,554 |
|  |  | Dead | 789 |
|  | 8 | Alive | 47,406 |
|  |  | Dead | 782 |
|  | 9 | Alive | 45,757 |
|  |  | Dead | 762 |
|  | 10 (least deprived) | Alive | 45,246 |
|  |  | Dead | 775 |
| **6** | 1 (most deprived) | Alive | 50,759 |
|  |  | Dead | 858 |
|  | 2 | Alive | 46,439 |
|  |  | Dead | 798 |
|  | 3 | Alive | 44,926 |
|  |  | Dead | 801 |
|  | 4 | Alive | 43,554 |
|  |  | Dead | 826 |
|  | 5 | Alive | 42,114 |
|  |  | Dead | 820 |
|  | 6 | Alive | 41,236 |
|  |  | Dead | 833 |
|  | 7 | Alive | 39,125 |
|  |  | Dead | 883 |
|  | 8 | Alive | 38,879 |
|  |  | Dead | 843 |
|  | 9 | Alive | 37,088 |
|  |  | Dead | 830 |
|  | 10 (least deprived) | Alive | 36,863 |
|  |  | Dead | 736 |
| **7** | 1 (most deprived) | Alive | 36,060 |
|  |  | Dead | 740 |
|  | 2 | Alive | 33,184 |
|  |  | Dead | 679 |
|  | 3 | Alive | 31,700 |
|  |  | Dead | 691 |
|  | 4 | Alive | 30,972 |
|  |  | Dead | 747 |
|  | 5 | Alive | 29,457 |
|  |  | Dead | 776 |
|  | 6 | Alive | 29,177 |
|  |  | Dead | 748 |
|  | 7 | Alive | 27,974 |
|  |  | Dead | 760 |
|  | 8 | Alive | 27,675 |
|  |  | Dead | 732 |
|  | 9 | Alive | 26,403 |
|  |  | Dead | 736 |
|  | 10 (least deprived) | Alive | 25,519 |
|  |  | Dead | 637 |
| **8** | 1 (most deprived) | Alive | 26,621 |
|  |  | Dead | 659 |
|  | 2 | Alive | 23,925 |
|  |  | Dead | 623 |
|  | 3 | Alive | 23,298 |
|  |  | Dead | 645 |
|  | 4 | Alive | 22,890 |
|  |  | Dead | 675 |
|  | 5 | Alive | 21,751 |
|  |  | Dead | 657 |
|  | 6 | Alive | 21,194 |
|  |  | Dead | 671 |
|  | 7 | Alive | 20,451 |
|  |  | Dead | 702 |
|  | 8 | Alive | 20,405 |
|  |  | Dead | 650 |
|  | 9 | Alive | 19,531 |
|  |  | Dead | 592 |
|  | 10 (least deprived) | Alive | 18,426 |
|  |  | Dead | 629 |
| **9** | 1 (most deprived) | Alive | 19,074 |
|  |  | Dead | 518 |
|  | 2 | Alive | 17,021 |
|  |  | Dead | 525 |
|  | 3 | Alive | 16,515 |
|  |  | Dead | 554 |
|  | 4 | Alive | 15,972 |
|  |  | Dead | 562 |
|  | 5 | Alive | 15,388 |
|  |  | Dead | 538 |
|  | 6 | Alive | 15,055 |
|  |  | Dead | 569 |
|  | 7 | Alive | 14,580 |
|  |  | Dead | 588 |
|  | 8 | Alive | 14,172 |
|  |  | Dead | 524 |
|  | 9 | Alive | 13,637 |
|  |  | Dead | 523 |
|  | 10 (least deprived) | Alive | 13,174 |
|  |  | Dead | 476 |
| **10** | 1 (most deprived) | Alive | 14,311 |
|  |  | Dead | 483 |
|  | 2 | Alive | 12,717 |
|  |  | Dead | 433 |
|  | 3 | Alive | 12,477 |
|  |  | Dead | 425 |
|  | 4 | Alive | 11,939 |
|  |  | Dead | 453 |
|  | 5 | Alive | 11,401 |
|  |  | Dead | 487 |
|  | 6 | Alive | 11,181 |
|  |  | Dead | 492 |
|  | 7 | Alive | 10,978 |
|  |  | Dead | 440 |
|  | 8 | Alive | 10,599 |
|  |  | Dead | 459 |
|  | 9 | Alive | 10,412 |
|  |  | Dead | 444 |
|  | 10 (least deprived) | Alive | 9,543 |
|  |  | Dead | 458 |
| **11** | 1 (most deprived) | Alive | 10,422 |
|  |  | Dead | 449 |
|  | 2 | Alive | 9,314 |
|  |  | Dead | 366 |
|  | 3 | Alive | 8,794 |
|  |  | Dead | 405 |
|  | 4 | Alive | 8,637 |
|  |  | Dead | 373 |
|  | 5 | Alive | 8,443 |
|  |  | Dead | 372 |
|  | 6 | Alive | 8,233 |
|  |  | Dead | 417 |
|  | 7 | Alive | 7,902 |
|  |  | Dead | 399 |
|  | 8 | Alive | 7,818 |
|  |  | Dead | 355 |
|  | 9 | Alive | 7,489 |
|  |  | Dead | 369 |
|  | 10 (least deprived) | Alive | 7,007 |
|  |  | Dead | 347 |
| **12** | 1 (most deprived) | Alive | 8,009 |
|  |  | Dead | 410 |
|  | 2 | Alive | 7,140 |
|  |  | Dead | 320 |
|  | 3 | Alive | 6,958 |
|  |  | Dead | 340 |
|  | 4 | Alive | 6,593 |
|  |  | Dead | 350 |
|  | 5 | Alive | 6,454 |
|  |  | Dead | 341 |
|  | 6 | Alive | 6,408 |
|  |  | Dead | 347 |
|  | 7 | Alive | 6,248 |
|  |  | Dead | 358 |
|  | 8 | Alive | 6,221 |
|  |  | Dead | 338 |
|  | 9 | Alive | 5,914 |
|  |  | Dead | 344 |
|  | 10 (least deprived) | Alive | 5,412 |
|  |  | Dead | 299 |
| **13 to 48** | 1 (most deprived) | Alive | 32,823 |
|  |  | Dead | 1,907 |
|  | 2 | Alive | 28,286 |
|  |  | Dead | 1,600 |
|  | 3 | Alive | 27,549 |
|  |  | Dead | 1,681 |
|  | 4 | Alive | 27,592 |
|  |  | Dead | 1,679 |
|  | 5 | Alive | 26,779 |
|  |  | Dead | 1,612 |
|  | 6 | Alive | 26,310 |
|  |  | Dead | 1,735 |
|  | 7 | Alive | 26,272 |
|  |  | Dead | 1,777 |
|  | 8 | Alive | 26,076 |
|  |  | Dead | 1,774 |
|  | 9 | Alive | 24,411 |
|  |  | Dead | 1,602 |
|  | 10 (least deprived) | Alive | 21,083 |
|  |  | Dead | 1,340 |

Notes:

Total time spent in the ED >12 hours has been aggregated due to low counts.

**Supplementary Table 9. Number of patients that were alive or dead within 30 days of leaving the ED, by total time spent in the ED and chief complaint**

| **Total time spent in the ED (hours, rounded to the nearest hour)** | **Chief complaint** | **Alive or dead within 30 days of discharge** | **Sample Size** |
| --- | --- | --- | --- |
| **0 to 3** | General / minor / admin | Alive | 239,903 |
|  |  | Dead | 1,907 |
|  | Airway / breathing | Alive | 181,114 |
|  |  | Dead | 2,729 |
|  | Circulation / chest | Alive | 302,680 |
|  |  | Dead | 1,395 |
|  | Environmental | Alive | 22,844 |
|  |  | Dead | 33 |
|  | Eye | Alive | 100,266 |
|  |  | Dead | 27 |
|  | Gastrointestinal | Alive | 277,367 |
|  |  | Dead | 1,249 |
|  | Genitourinary | Alive | 85,295 |
|  |  | Dead | 339 |
|  | Head and neck | Alive | 182,702 |
|  |  | Dead | 113 |
|  | Neurological | Alive | 194,094 |
|  |  | Dead | 2,469 |
|  | Obstetrics and gynaecology | Alive | 54,794 |
|  |  | Dead | 17 |
|  | Psychosocial / Behaviour change | Alive | 51,902 |
|  |  | Dead | 94 |
|  | Skin | Alive | 267,628 |
|  |  | Dead | 164 |
|  | Trauma / musculoskeletal | Alive | 1,284,244 |
|  |  | Dead | 1,090 |
| **4 and 5** | General / minor / admin | Alive | 146,292 |
|  |  | Dead | 3,387 |
|  | Airway / breathing | Alive | 148,444 |
|  |  | Dead | 5,609 |
|  | Circulation / chest | Alive | 260,957 |
|  |  | Dead | 2,034 |
|  | Environmental | Alive | 12,997 |
|  |  | Dead | 59 |
|  | Eye | Alive | 26,521 |
|  |  | Dead | 26 |
|  | Gastrointestinal | Alive | 245,239 |
|  |  | Dead | 2,822 |
|  | Genitourinary | Alive | 58,837 |
|  |  | Dead | 620 |
|  | Head and neck | Alive | 64,787 |
|  |  | Dead | 160 |
|  | Neurological | Alive | 187,102 |
|  |  | Dead | 4,032 |
|  | Obstetrics and gynaecology | Alive | 27,761 |
|  |  | Dead | 18 |
|  | Psychosocial / Behaviour change | Alive | 43,789 |
|  |  | Dead | 146 |
|  | Skin | Alive | 82,595 |
|  |  | Dead | 209 |
|  | Trauma / musculoskeletal | Alive | 458,029 |
|  |  | Dead | 1,975 |
| **6 and 7** | General / minor / admin | Alive | 58,347 |
|  |  | Dead | 2,376 |
|  | Airway / breathing | Alive | 68,099 |
|  |  | Dead | 4,250 |
|  | Circulation / chest | Alive | 119,592 |
|  |  | Dead | 1,405 |
|  | Environmental | Alive | 5,224 |
|  |  | Dead | 43 |
|  | Eye | Alive | 7,251 |
|  |  | Dead | 20 |
|  | Gastrointestinal | Alive | 113,273 |
|  |  | Dead | 2,157 |
|  | Genitourinary | Alive | 24,985 |
|  |  | Dead | 423 |
|  | Head and neck | Alive | 20,664 |
|  |  | Dead | 115 |
|  | Neurological | Alive | 92,762 |
|  |  | Dead | 2,863 |
|  | Obstetrics and gynaecology | Alive | 9,422 |
|  |  | Dead | 13 |
|  | Psychosocial / Behaviour change | Alive | 21,246 |
|  |  | Dead | 72 |
|  | Skin | Alive | 24,384 |
|  |  | Dead | 170 |
|  | Trauma / musculoskeletal | Alive | 153,855 |
|  |  | Dead | 1,567 |
| **8 to 11** | General / minor / admin | Alive | 48,707 |
|  |  | Dead | 3,250 |
|  | Airway / breathing | Alive | 61,863 |
|  |  | Dead | 5,658 |
|  | Circulation / chest | Alive | 95,152 |
|  |  | Dead | 1,797 |
|  | Environmental | Alive | 3,684 |
|  |  | Dead | 58 |
|  | Eye | Alive | 3,987 |
|  |  | Dead | 16 |
|  | Gastrointestinal | Alive | 96,161 |
|  |  | Dead | 2,916 |
|  | Genitourinary | Alive | 19,825 |
|  |  | Dead | 485 |
|  | Head and neck | Alive | 13,118 |
|  |  | Dead | 148 |
|  | Neurological | Alive | 84,549 |
|  |  | Dead | 3,732 |
|  | Obstetrics and gynaecology | Alive | 5,635 |
|  |  | Dead | 28 |
|  | Psychosocial / Behaviour change | Alive | 17,245 |
|  |  | Dead | 88 |
|  | Skin | Alive | 14,188 |
|  |  | Dead | 169 |
|  | Trauma / musculoskeletal | Alive | 108,583 |
|  |  | Dead | 1,961 |
| **12 to 48** | General / minor / admin | Alive | 35,243 |
|  |  | Dead | 3,402 |
|  | Airway / breathing | Alive | 44,247 |
|  |  | Dead | 5,341 |
|  | Circulation / chest | Alive | 50,733 |
|  |  | Dead | 1,689 |
|  | Environmental | Alive | 2,024 |
|  |  | Dead | 62 |
|  | Eye | Alive | 1,211 |
|  |  | Dead | 12 |
|  | Gastrointestinal | Alive | 52,327 |
|  |  | Dead | 2,945 |
|  | Genitourinary | Alive | 10,446 |
|  |  | Dead | 551 |
|  | Head and neck | Alive | 5,147 |
|  |  | Dead | 148 |
|  | Neurological | Alive | 58,498 |
|  |  | Dead | 3,841 |
|  | Obstetrics and gynaecology | Alive | 1,659 |
|  |  | Dead | 15 |
|  | Psychosocial / Behaviour change | Alive | 12,910 |
|  |  | Dead | 127 |
|  | Skin | Alive | 5,424 |
|  |  | Dead | 171 |
|  | Trauma / musculoskeletal | Alive | 52,669 |
|  |  | Dead | 1,850 |

Notes:

Total time spent in the ED has been aggregated into groups due to low counts.

**Supplementary Table 10. Number of patients that were alive or dead within 30 days of leaving the ED, by total time spent in the ED and acuity level**

| **Total time spent in the ED (hours, rounded to the nearest hour)** | **Acuity level** | **Alive or dead within 30 days of discharge** | **Sample Size** |
| --- | --- | --- | --- |
| **0hr to 59m** | Very urgent | Alive | 4,194 |
|  |  | Dead | 208 |
|  | Urgent | Alive | 32,753 |
|  |  | Dead | 147 |
|  | Standard | Alive | 91,586 |
|  |  | Dead | 124 |
|  | Low | Alive | 27,897 |
|  |  | Dead | 21 |
| **1** | Very urgent | Alive | 27,836 |
|  |  | Dead | 417 |
|  | Urgent | Alive | 158,332 |
|  |  | Dead | 424 |
|  | Standard | Alive | 478,085 |
|  |  | Dead | 364 |
|  | Low | Alive | 50,113 |
|  |  | Dead | 50 |
| **2** | Very urgent | Alive | 63,762 |
|  |  | Dead | 1,159 |
|  | Urgent | Alive | 306,739 |
|  |  | Dead | 1,147 |
|  | Standard | Alive | 670,116 |
|  |  | Dead | 579 |
|  | Low | Alive | 52,148 |
|  |  | Dead | 74 |
| **3** | Very urgent | Alive | 105,144 |
|  |  | Dead | 2,475 |
|  | Urgent | Alive | 469,833 |
|  |  | Dead | 3,229 |
|  | Standard | Alive | 656,984 |
|  |  | Dead | 1,078 |
|  | Low | Alive | 49,311 |
|  |  | Dead | 130 |
| **4** | Very urgent | Alive | 130,128 |
|  |  | Dead | 4,302 |
|  | Urgent | Alive | 554,469 |
|  |  | Dead | 6,672 |
|  | Standard | Alive | 521,145 |
|  |  | Dead | 2,122 |
|  | Low | Alive | 41,517 |
|  |  | Dead | 234 |
| **5** | Very urgent | Alive | 58,119 |
|  |  | Dead | 2,520 |
|  | Urgent | Alive | 244,834 |
|  |  | Dead | 3,925 |
|  | Standard | Alive | 193,132 |
|  |  | Dead | 1,163 |
|  | Low | Alive | 20,006 |
|  |  | Dead | 159 |
| **6** | Very urgent | Alive | 51,757 |
|  |  | Dead | 2,555 |
|  | Urgent | Alive | 211,925 |
|  |  | Dead | 4,231 |
|  | Standard | Alive | 141,595 |
|  |  | Dead | 1,275 |
|  | Low | Alive | 15,706 |
|  |  | Dead | 167 |
| **7** | Very urgent | Alive | 38,617 |
|  |  | Dead | 2,284 |
|  | Urgent | Alive | 154,921 |
|  |  | Dead | 3,641 |
|  | Standard | Alive | 93,628 |
|  |  | Dead | 1,151 |
|  | Low | Alive | 10,955 |
|  |  | Dead | 170 |
| **8** | Very urgent | Alive | 29,693 |
|  |  | Dead | 1,926 |
|  | Urgent | Alive | 115,981 |
|  |  | Dead | 3,359 |
|  | Standard | Alive | 64,861 |
|  |  | Dead | 1,024 |
|  | Low | Alive | 7,957 |
|  |  | Dead | 194 |
| **9** | Very urgent | Alive | 21,811 |
|  |  | Dead | 1,583 |
|  | Urgent | Alive | 83,550 |
|  |  | Dead | 2,843 |
|  | Standard | Alive | 43,604 |
|  |  | Dead | 800 |
|  | Low | Alive | 5,623 |
|  |  | Dead | 151 |
| **10** | Very urgent | Alive | 17,022 |
|  |  | Dead | 1,360 |
|  | Urgent | Alive | 63,576 |
|  |  | Dead | 2,416 |
|  | Standard | Alive | 30,789 |
|  |  | Dead | 682 |
|  | Low | Alive | 4,171 |
|  |  | Dead | 116 |
| **11** | Very urgent | Alive | 13,022 |
|  |  | Dead | 1,136 |
|  | Urgent | Alive | 46,944 |
|  |  | Dead | 1,997 |
|  | Standard | Alive | 21,252 |
|  |  | Dead | 591 |
|  | Low | Alive | 2,841 |
|  |  | Dead | 128 |
| **12** | Very urgent | Alive | 10,859 |
|  |  | Dead | 996 |
|  | Urgent | Alive | 36,847 |
|  |  | Dead | 1,844 |
|  | Standard | Alive | 15,423 |
|  |  | Dead | 505 |
|  | Low | Alive | 2,228 |
|  |  | Dead | 102 |
| **13 to 48** | Very urgent | Alive | 47,635 |
|  |  | Dead | 4,668 |
|  | Urgent | Alive | 150,444 |
|  |  | Dead | 8,882 |
|  | Standard | Alive | 56,661 |
|  |  | Dead | 2,425 |
|  | Low | Alive | 12,441 |
|  |  | Dead | 732 |

Notes:

Total time spent in the ED >12 hours has been aggregated due to low counts.

**Supplementary Figure 1. Estimated marginal probability of all-cause, 30-day mortality as a function of time spent in the ED, for individuals in the ‘immediate’ acuity group**

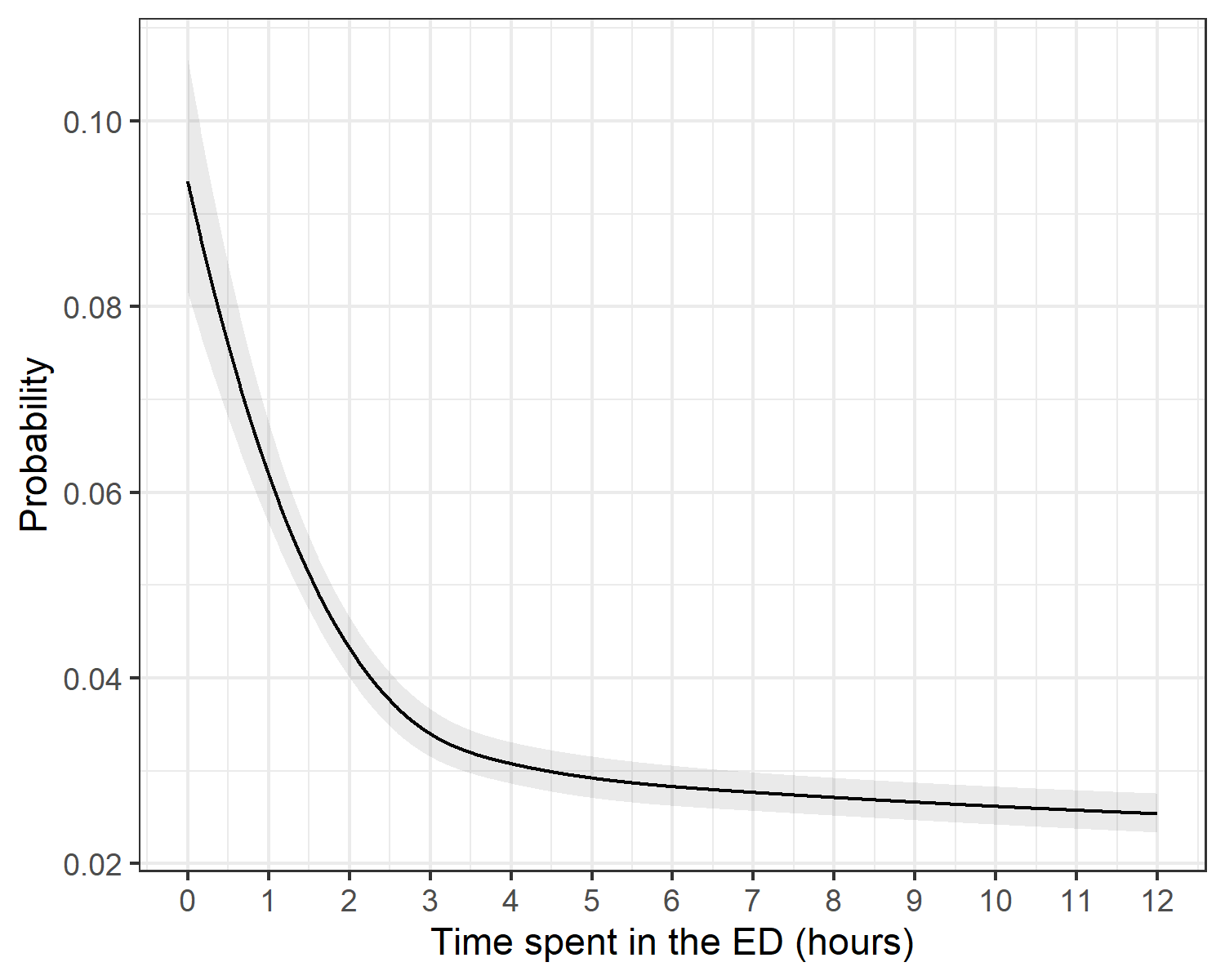

Patients in the “immediate” acuity group were excluded from the analysis because of the high mortality rate within a short time after arriving at the ED in this group. Analysis conducted on this group has been included below.

In our study period, 124,078 individuals attended the ED, had a recorded acuity level of ‘Immediate’ and met our study criteria.

Supplementary Figure 2 shows the estimated marginal probabilities of 30-day mortality following the first visit to the ED against time spent in the ED (in hours) for those in the ‘immediate’ acuity group. It demonstrates an increased probability of 30-day mortality of 0.094 (95% confidence interval: 0.082 to 0.011) when the patient first arrives at the ED. This sharply declines from 0 to 3 hours spent in the ED and begins to plateau after approximately 3 hours. This decline in the probability of post-discharge mortality over the first 3 hours of time spent in the ED is likely to be the result of residual confounding by illness severity for patients in the ‘immediate’ acuity group (i.e. those requiring resuscitation being prioritised for immediate treatment), rather than a causal relationship.

**Supplementary Figure 2. Estimated marginal probability of all-cause, 30-day mortality as a function of time to first treatment in the ED, for individuals in the ‘Very Urgent’, ‘Urgent’, ‘Standard’ and ‘Low’ acuity groups**

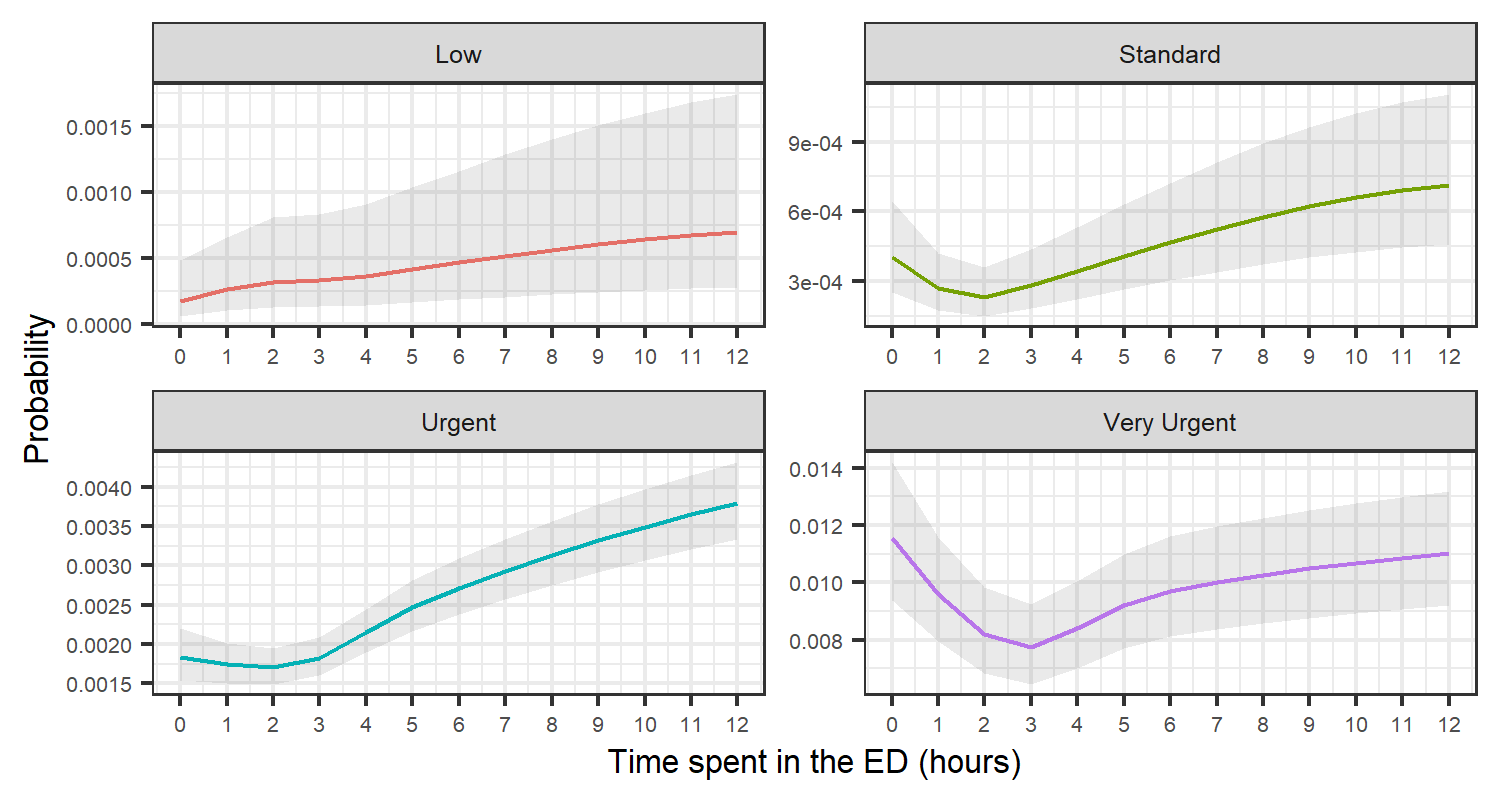

Notes:

1. The y-axis range is not consistent between the panels.
2. The numbers on the y-axis are in scientific notion. For example, ‘9e-04’ represents 9 x10^-4^, which is 0.0009. This notation is used to express very small or very large numbers compactly.

**Supplementary Figure 3. Sample flow diagram**

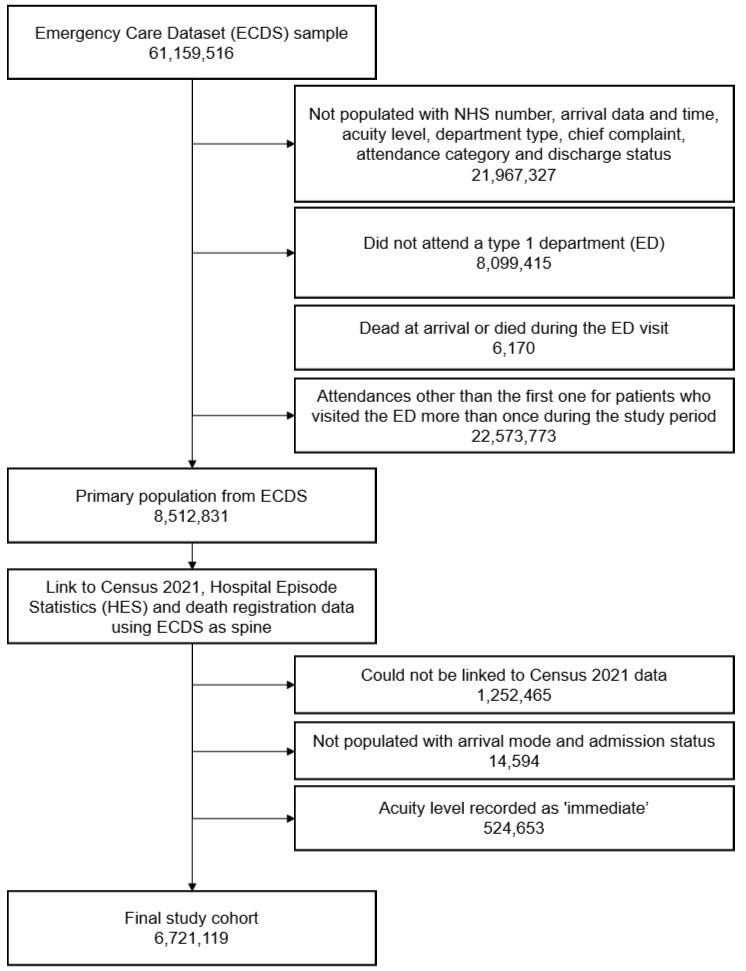

**Supplementary Figure 4. Distribution of total time spent in the ED up to 48 hours**

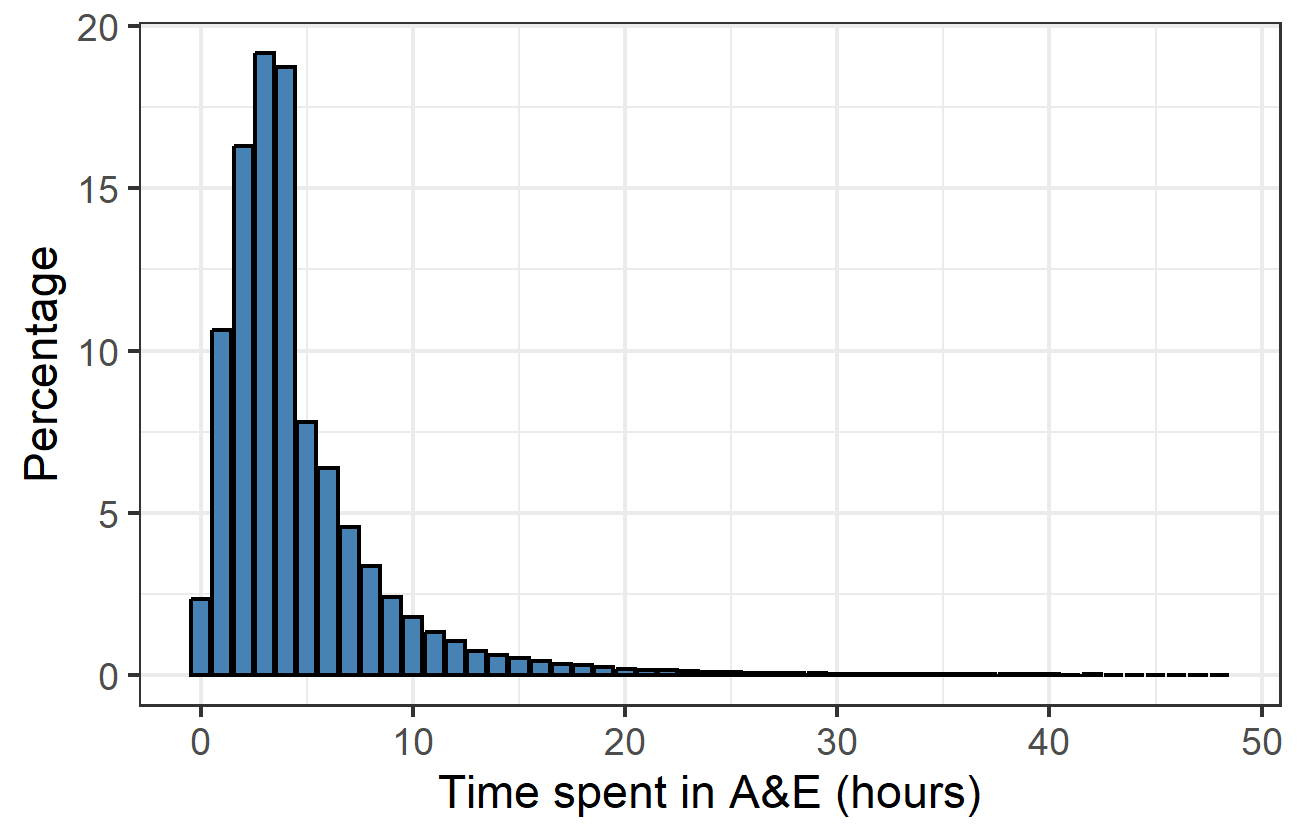

Time spent in the ED (hours)

Notes:

1. Time spent in the ED has been rounded to the nearest hour.

**Supplementary Figure 5. Adjusted odds ratios for all-cause, 30-day mortality as a function of time spent in the ED, compared to two hours**

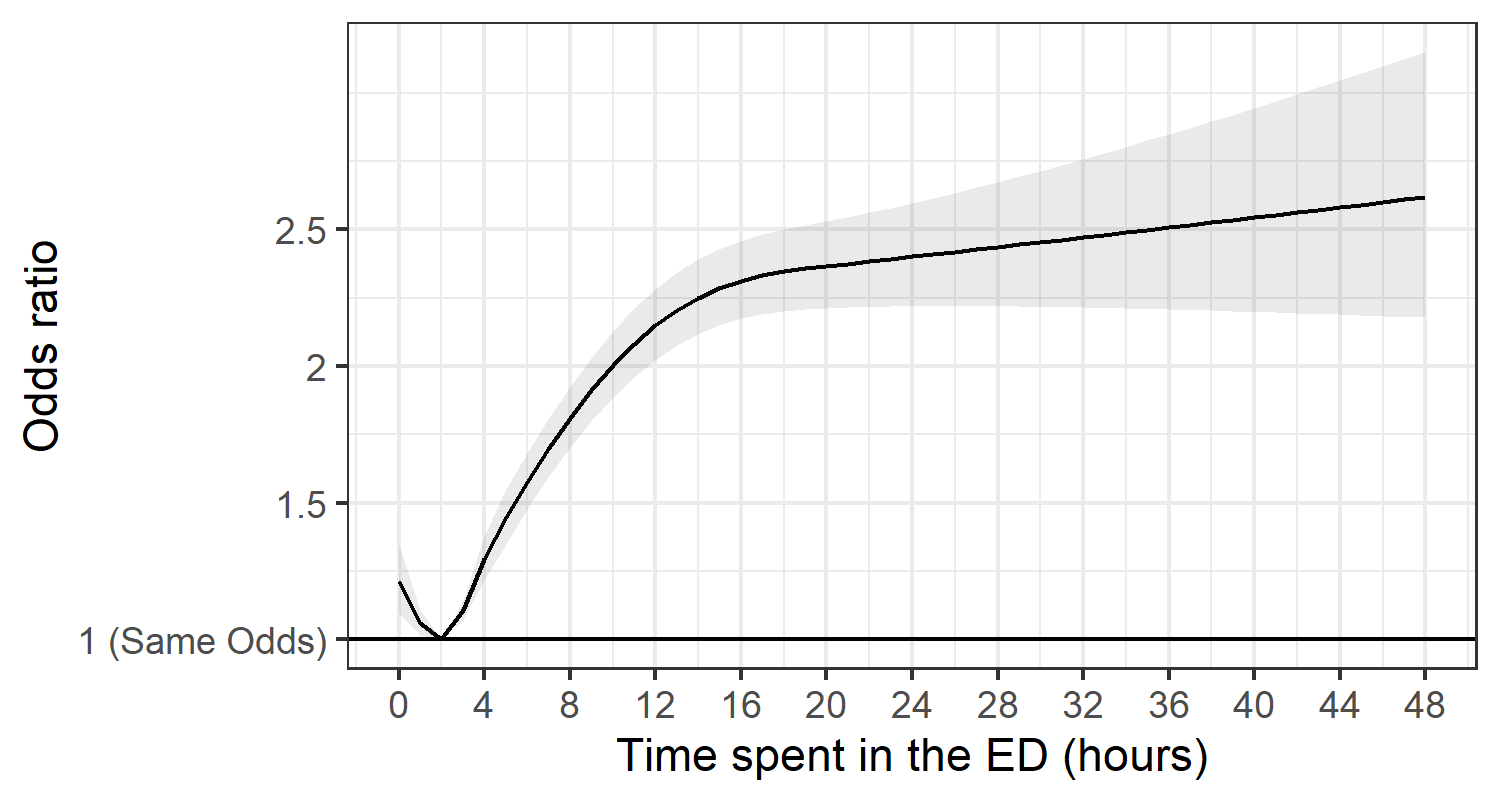

Notes:

1. The estimates are adjusted for the covariates outlined in the Methods section.
2. The shaded area represents the 95% confidence interval around the point estimates.

**Supplementary Figure 6.** **Adjusted odds ratios compared to two hours for all-cause, 30-day mortality as a function of time spent in the ED, by age group, up to 12 hours**

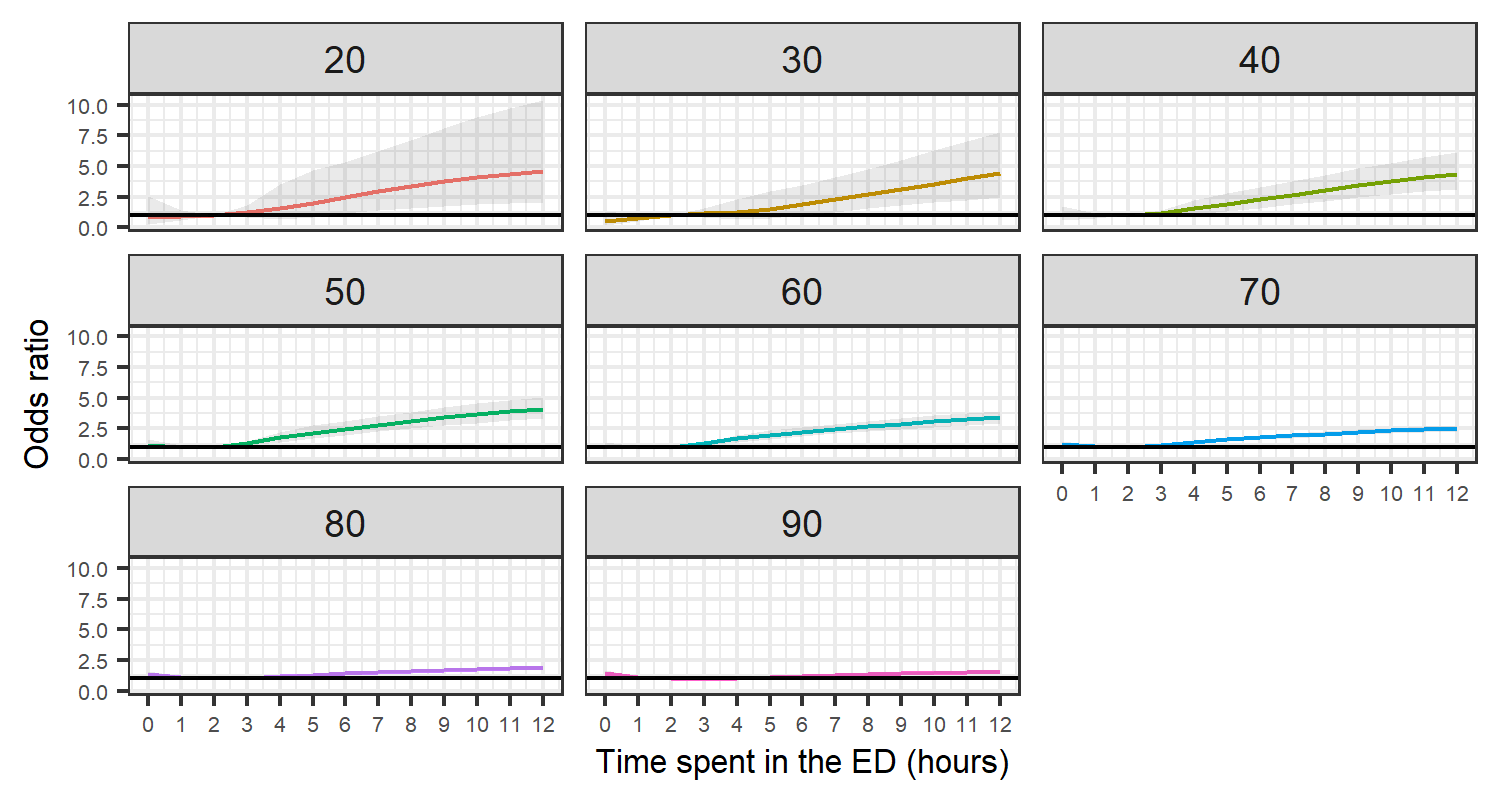

Notes:

1. The estimates are adjusted for the covariates outlined in the Methods section.
2. The shaded area represents the 95% confidence interval around the point estimates.
3. Each graph represents the odds ratios compared to two hours at specific ages

**Supplementary Figure 7 Adjusted odds ratios compared to two hours for all-cause, 30-day mortality as a function of time spent in the ED, by region of residence, up to 12 hours**

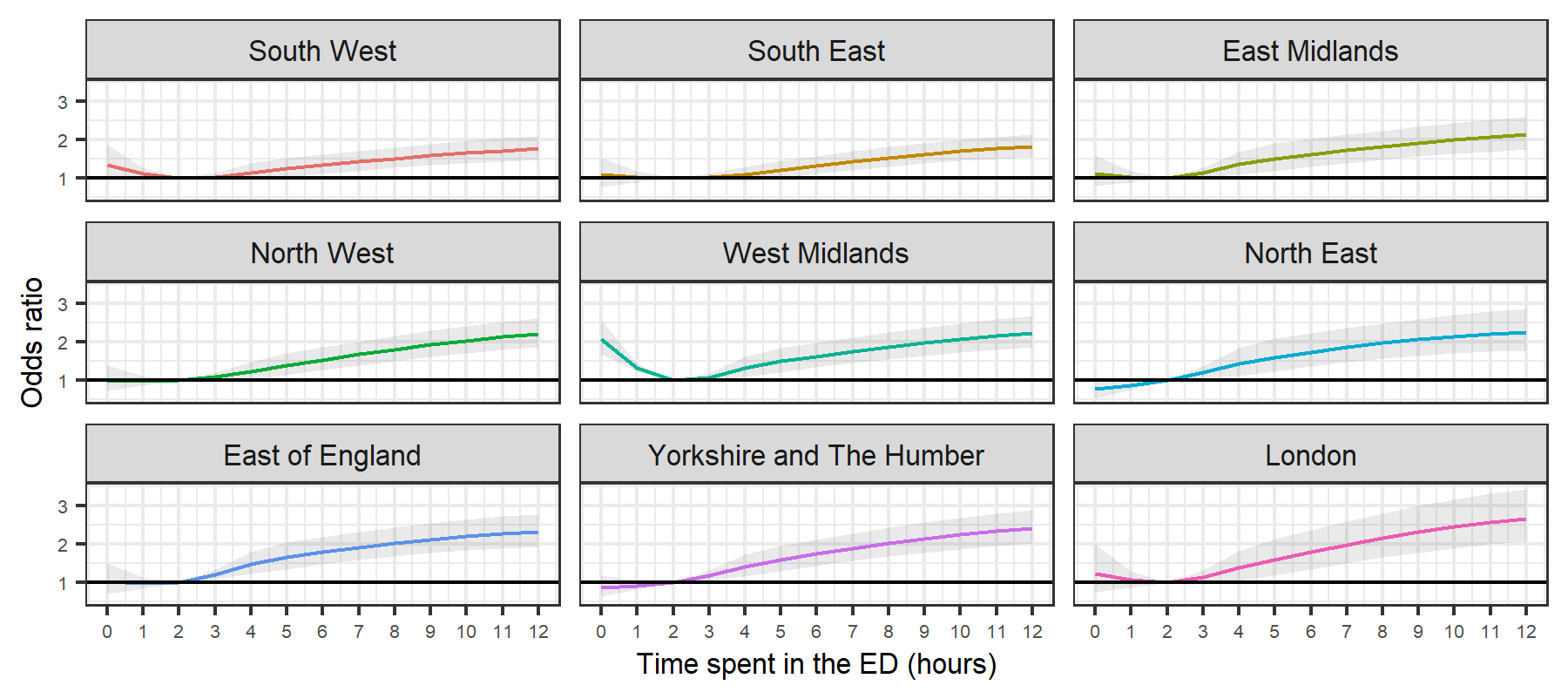

Notes:

1. The estimates are adjusted for the covariates outlined in the Methods section.
2. The shaded area represents the 95% confidence interval around the point estimates.

**Supplementary Figure 8. Adjusted odds ratios compared to two hours for all-cause, 30-day mortality as a function of time spent in the ED, by relative area deprivation decile group, up to 12 hours**

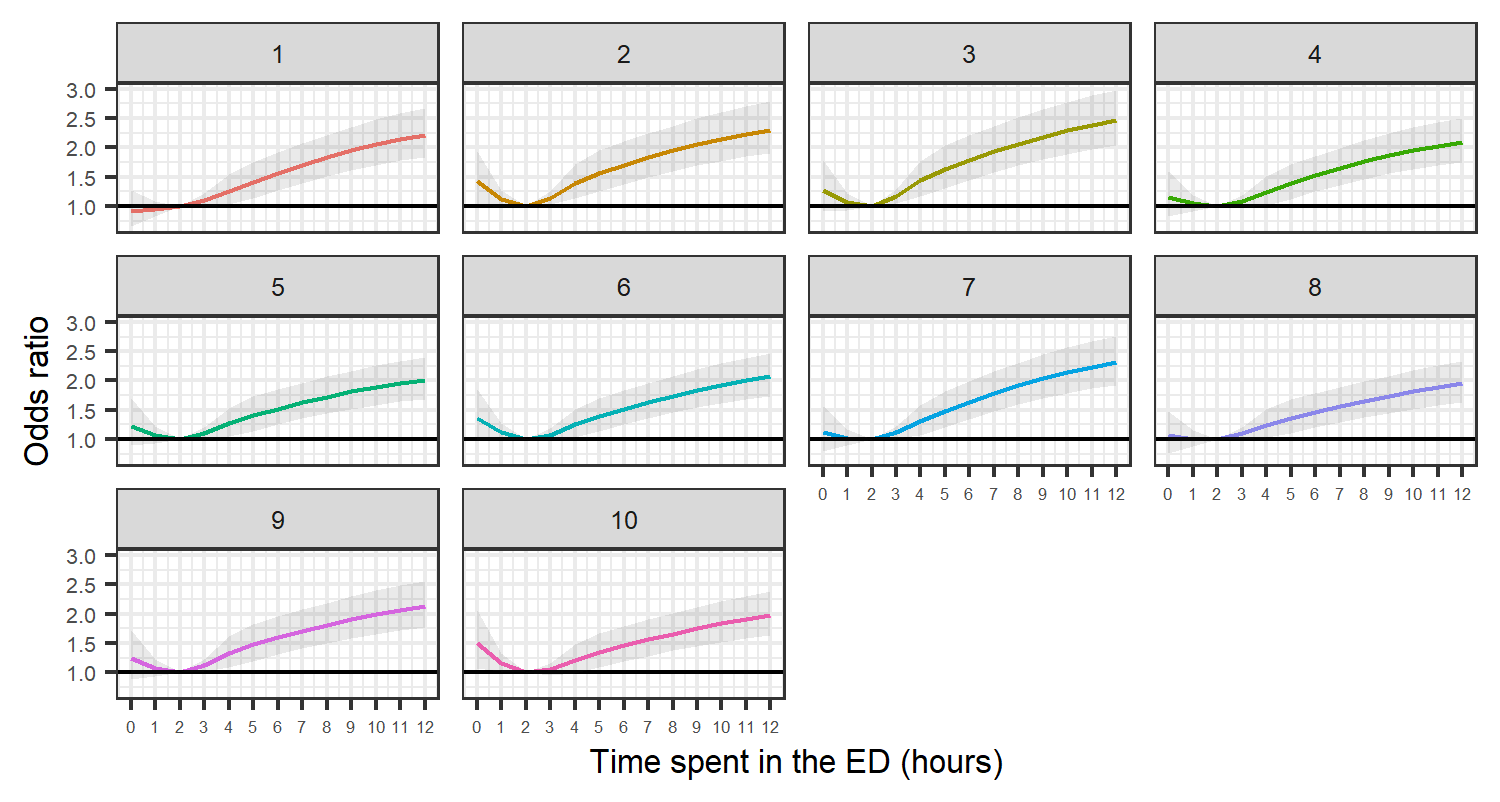

Notes:

1. The estimates are adjusted for the covariates outlined in the Methods section.
2. The shaded area represents the 95% confidence interval around the point estimates.

**Supplementary Figure 9. Adjusted odds ratios compared to two hours for all-cause, 30-day mortality as a function of time spent in the ED, additionally adjusted for admission status – sensitivity analysis including admission status as a covariate in the model**

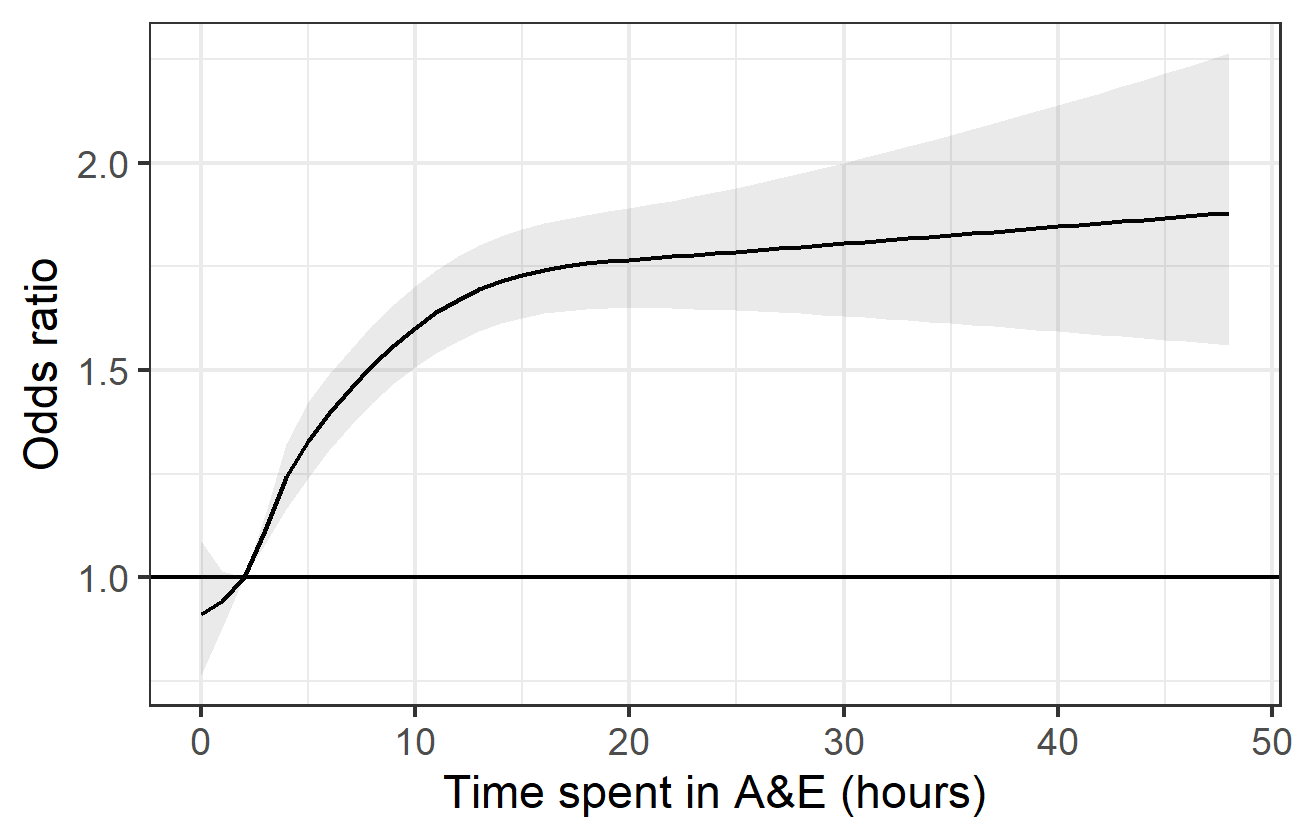

Time spent in the ED (hours)

Notes:

1. The estimates are adjusted for the covariates outlined in the Methods section.
2. The shaded area represents the 95% confidence interval around the point estimates.
